# Simulation-informed evaluation of microvascular parameter mapping for diffusion MR imaging of solid tumours

**DOI:** 10.1101/2025.08.27.25334553

**Authors:** Anna Kira Voronova, Olivia Prior, Athanasios Grigoriou, Francesc Salvà, Elena Elez, Luz M. Atlagich, Roser Sala-Llonch, Marco Palombo, Els Fieremans, Dmitry S. Novikov, Raquel Perez-Lopez, Francesco Grussu

## Abstract

**Purpose:** We aim to inform the design of new diffusion MRI (dMRI) approaches for microvasculature mapping that enhance the biological specificity of imaging towards cancer.

**Methods:** We adopted simulation-informed modelling of the vascular dMRI signal. We synthesised signals from 1500 synthetic vascular networks, for a variety of protocols (flow-compensated (FC), non-compensated (NC), hybrid), featuring different *b* samplings and diffusion times. We estimated the number of independent, recoverable signal degrees of freedom in presence of noise (signal-to-noise ratio of 5), and ranked 12 microvascular metrics depending on the quality of their estimation. Lastly, we demonstrated the feasibility of estimating the top-ranking metrics on 3T dMRI of a healthy volunteer and of a metastatic colorectal cancer (CRC) patient.

**Results:** Both NC and FC synthetic vascular signals exhibit complex behaviour, e.g., non-zero kurtosis and diffusion time dependence. Two independent degrees of freedom appear recoverable from directionally-averaged vascular signals (SNR of 5). Mean volumetric flow rate *q*_*m*_ and an *Apparent Network Branching* (ANB) index maximise correlations between ground truth and estimated values *in silico*. Their estimation is proposed for *in vivo* imaging, and demonstrated herein. In the patient, both *q*_*m*_ and *ANB* detect re-vascularisation after 3 months of targeted therapy against liver metastases, consistently with *Intra-Voxel Incoherent Motion* (IVIM) metrics.

**Conclusions:** Simulation-based modelling of the vascular dMRI signal informs the design of promising approaches for *in vivo* microvasculature characterisation.

## 1 Introduction

Motion-probing gradients are used in diffusion Magnetic Resonance Imaging (dMRI) to sensitise the MRI signal to different types of spin motion[1], including flow from microperfusion, without relying on contrast agents[2, 3, 4]. An example of a well-established method for capillary flow quantification is *intra-voxel incoherent motion* (IVIM) dMRI [4, 5, 6, 7], a common approach for voxel-wise estimation of the apparent perfusion fraction (*f*_*v*_) and of the pseudo-diffusion coefficient (*D*^*^)[8, 9]. IVIM indices offer promise in liver malignancy detection [10] or hyper-vascularisation assessment [11], and correlate with vessel histology[12, 13]. However, despite these encouraging data, IVIM metrics have so far failed to make a lasting impact in the clinic, being semi-quantitative and protocol-dependent [14], facts that hinder their clinical adoption.

Recent research has focussed on the design of new signal representations that enhance the biological specificity of dMRI beyond IVIM[15, 4, 8, 9, 16, 17, 18]. In this context, the numerical simulation of blood flow within synthetic vascular networks has shown promise as a way to increase the realism of vascular dMRI signal models, potentially paving the way to a new generation of techniques[19, 20, 21, 22]. *In this work, we adopt this powerful approach and use simulation-based modelling to inform the development of new dMRI microvascular parameter mapping approaches*. In the long-term, our study aims to support the introduction of new quantitative vascular biomarkers for precision medicine, being these still sought, for example, in oncology.

We used capillary networks traced on histological liver tissue to simulate vascular dMRI signals for realistic flow-compensated (FC) and non-compensated (NC) acquisitions [23]. We analysed such signals to compute the number of recoverable, independent vascular parameters clinical signal-to-noise ratio (SNR), and investigated which microvascular properties can be practically retrieved through model fitting. Lastly, we tested the estimation of the most promising microvascular parameters *in vivo*, demonstrating the potential of physics-informed dMRI modelling for microvascular mapping.

## 2 Methods

The following sub-sections outline the details of our *in silico* and *in vivo* studies.

### 2.1 Vascular signal analysis *in silico*

#### 2.1.1 Microvascular networks

We used 15 freely available 2D vascular networks from a recent study[22], derived from liver biopsies (permanent address: https://github.com/radiomicsgroup/SpinFlowSim/tree/main/networks). The networks are characterised by a set of nodes, connected through capillary segments, and feature one inlet and one outlet. We generated 1500 unique networks through 100 tridimensional realisations of each 2D network (Supporting Information Figure S1). For each realisation, we perturbed the (*x, y*) position of each node, varied each segment diameter, and changed inlet/outlet. Additionally, we also simulated 3D depth using an exponential function, with maximum depth achieved in the network centre. Radii were perturbed of up to *±* 40% from the original value; *x*/*y* node positions of up to *±*2 *µ*m; and the network depth of *±z*_*max*_ = 150 *µ*m. Moreover, for each perturbation we also removed 3% of the capillary segments. All perturbations were drawn from the uniform distribution.

We simulated an input volumetric flow rate (VFR) *q*_*in*_ in 10 equally-distributed values in [1.5 *·*10^−4^; 2.75 *·*10^−3^] mm^3^*/*s, and obtained per-segment VFR *q* and mean velocity vector **v**. Each network instantiation was seeded uniformly with 5000 spins. The *n*-th spin trajectory **p**_*n*_(*t*) was obtained via

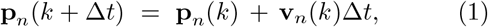

for *k* = {0, Δ*t*, 2Δ*t*, …}. Above, Δ*t* is the temporal resolution (set to 10 *µ*s), and **v**_*n*_(*t*) is the instantaneous spin velocity. Simulations were based on SpinFlowSim[22] (https://github.com/radiomicsgroup/SpinFlowSim).

We characterised each network realisation by computing a wide range of properties, namely: statistics of the distributions of the VFR *q*, velocity *v*, radius *r*, and capillary length *L*; indices describing its size, connectivity and complexity.

Regarding *v* and *q*, we computed mean and standard deviation across all capillary segments, i.e.,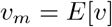 and 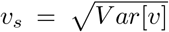 and 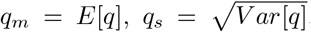.We also computed the path-weighted mean velocity 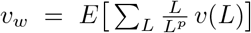 and the path-weighted mean 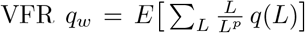. This was done by averaging over all possible input/output paths a weighted velocity/VFR index. This index is obtained by averaging *v* (or *q*) across capillaries, weighting the average with the ratio 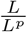,where *L*^*p*^ = ∑ *L* is the total path length.

We obtained similar indices of mean and path-weighted mean for the capillary radius, i.e.: *r*_*m*_ = *E*[*r*], We also computed (i) the mean capillary segment and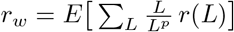.

We also computed (i) the mean capillary segment length *L*_*m*_, (ii) the mean input/output path length

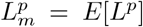, (iii) the number of input/output (IO)paths *N*_*paths*_, and (iv) the *apparent network branching* (*ANB*)[22]. *ANB* estimates the average number of capillary segments spins travel through during a reference time of 100 ms.

#### 2.1.3 dMRI signal synthesis

Motion-sensitising gradient waveforms **G**(*t*) were used to obtain magnitude dMRI signals as[24]

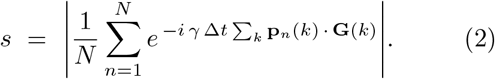

We used NC pulsed-gradient spin echo (PGSE) monopolar waveforms[25] and FC bipolar waveforms[26], compensating for velocity. Both NC and FC gradients were linearly polarised[27], and both refocus stationary spins, since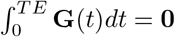. FC waveforms also refocus spins flowing at constant velocity (ballistic regime[9]), since they null the 1^st^ gradient moment 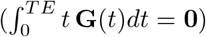. NC waveforms were parametrised by the diffusion gradient duration/separation/strength *δ*/Δ/*G*, while FC waveforms by *G*, Δ and by the oscillation half-period *τ* (Supporting Information Figure S2). Note that *b* = *γ*^2^*G*^2^*δ*^2^(Δ− *δ/*3) for the NC waveforms, and 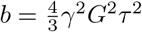 for the FC ones.

We synthesised signals for several different protocols, using 15 uniformly-distributed gradient directions[28] for each *b*. The protocols were:

- a *NC protocol*, with b-values matching the *in vivo* acquisitions (see below), i.e., *b* = {0, 10, 20, 40, 70, 100} s*/*mm^2^. We used a fixed Δ of either 30 ms or 50 ms, and gradient duration *δ* = 6 ms. Signals from the 15 directions were averaged.
- A *FC protocol*, with the same 6 b-values, as above. We fixed Δ to 30 ms, and used a half-period *τ* of either 3 ms or 10 ms. Signals from the 15 directions were averaged.
- A *hybrid protocol*, obtained by alternating NC and FC b-values from the two protocols above.
- A *richNC protocol*, featuring NC PGSE measurements as above (Δ = 30 ms, *δ* = 6 ms), but a richer b-value sampling (20 b-values in the range [0; 100] s/*mm*^2^). Two versions of the protocol were obtained: one with directional averaging at fixed *b*, and one without averaging.
- A *richFC protocol*, featuring FC measurements as above (Δ = 30 ms, *τ* = 10 ms), but again, a richer*b* sampling (20 values of *b* in [0; 100] s/*mm*^2^), and again with/without directional averaging.
- A *hybrid rich protocol*, obtained by alternating richNC and richFC b-values from the two protocols above.

We also visualised dMRI signals for a wide range of diffusion times, i.e., Δ = {10, 50, 100} ms for both FC and NC protocols (with *δ* = 0.5 ms for the NC protocol and *τ* = 1 ms for the FC one).

Signals were corrupted with Rician noise (SNR of 5 and 20 at *b* = 0).

#### 2.1.3 Analysis: degrees of freedom estimation

We performed Singular Value Decomposition (SVD) of matrices storing noisy and noise-free signals to estimate the number of independent, detectable microvascular degrees of freedom[29, 30] *N*_*p*_. We stacked all *Q* = 1500 microvascular signals in matrices of size *Q ×M*, where *M* is the number of protocol measurements, and computed the *min*(*Q, M*) independent SVs. For each protocol and SNR, we compared noisy/noise-free SVs *λ* to estimate *N*_*p*_ by counting the noisy SVs such that

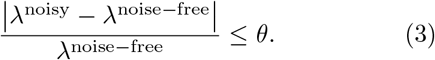

We used *θ* = 0.17, as this value roughly corresponded to the threshold provided by Marchenko-Pastur Principal Component Analysis (MP-PCA)[29, 30] for protocols featuring large *M* (e.g., rich protocols).

#### 2.1.4 Analysis: vascular property estimation and ranking

We investigated which microvascular properties can be best inferred from noisy measurements (Fig. 1). We trained radial basis function (RBF)[22] numerical forward models on noise-free signals from 14 out of 15 networks regressors. Afterwards, the trained RBF models were fit to noisy signals from the 15^th^ network through maximum-likelihood inference[31], in a leave-one-out fashion, estimating each vascular property in turn.

**Figure 1.**
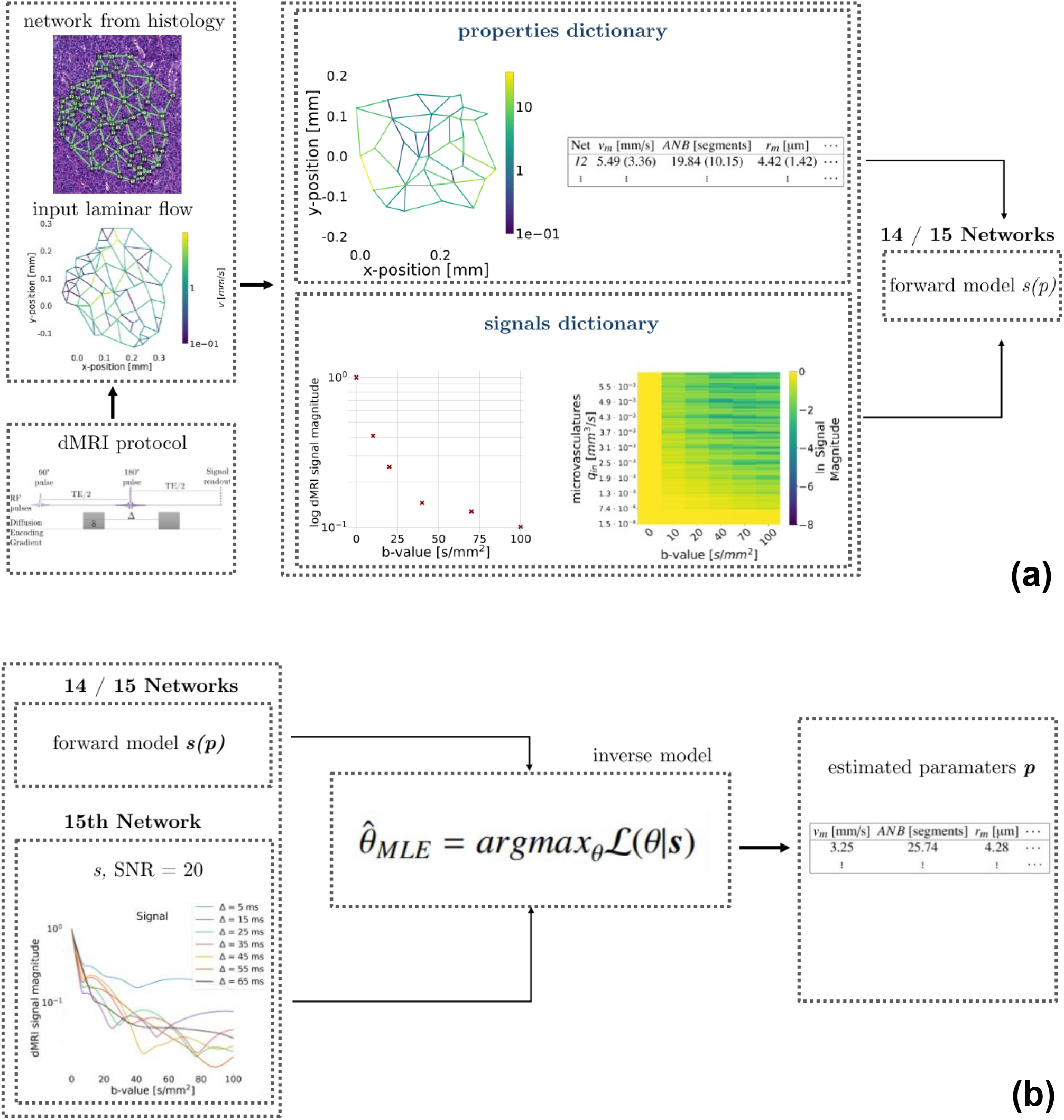
Illustration of the simulation-informed framework developed to study microvascular parameter inference *in silico*, which relies on 15 synthetic vascular networks available from a previous study. (a): firstly, synthetic dMRI signals for several acquisition protocols were generated using *SpinFlowSim*. Noise-free signal from 14 out of 15 networks in turn were used to build numerical forward models, predicting dMRI signals from a vascular property of interest. (b): afterwards, the numerical signal models were plugged into standard maximum-likelihood fitting routines, which estimated the vascular property of interest on noisy signals from the 15^th^ network, in a leave-one-out fashion.

We compared estimated and ground truth properties by computing the Spearman’s correlation coefficient *r*_*s*_, and a Bias Index (BI)[32]. Both were evaluated pooling together predictions for all 1500 networks across. Confidence intervals were obtained by recording ranges across folds. Properties were ranked according to decreasing *r*_*s*_.

#### 2.1.5 Analysis: relationship between vascular properties and vascular signal features

Finally, we investigated how the top-ranking metrics are encoded in the vascular dMRI signal, by assesing their relationship with the main dMRI signal cumulants, i.e., the apparent pseudo-diffusion and kurtosis coefficients, referred to as *D*^*^ and *K*^*^. These characterise the signal slope and curvature in log-scale as a function of the b-value, and were computed by fitting

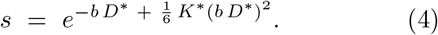

We computed the Spearman’s correlation coefficient *r*_*s*_ between the top-ranking vascular properties and *D*^*^/*K*^*^, and between the two top-ranking metrics themselves.

### 2.2 Vascular signal analysis *in vivo*

Lastly, we demonstrated the feasibility of estimating the top-ranking metrics from the *in silico* study on *in vivo* images. For this purpose, we scanned a male healthy volunteer (30-34 years old range) and a male patient (50-54 years old range) suffering from CRC, with liver metastases. The patient was scanned immediately before receiving targeted therapy based on Ecorafenib, Binimetinib and Cetuximab, and after 3 months of treatment. Informed written consent was obtained, and the study was approved by the Vall d’Hebron University Hospital Research Ethics Committee (Barcelona, Spain; CEIm PR(AG)362/2021, PR(IDI)109/2022).

#### 2.2.1 MRI acquisition and processing

Volunteers were scanned on a 3T GE SIGNA Pioneer scanner at abdominal level. The protocol included structural imaging and dMRI, with salient parameters: resolution 2.4 ×2.4 ×6 mm^3^, TE = 75 ms, TR = 12 s (respiratory-gated), bandwidth 3333 Hz/pixel, trace DW imaging; NEX = 2; ASSET = 2, *b* = {0, 10, 20, 40, 70, 100, 500, 1000, 1250, 1500} s*/*mm^2^, with gradient timings: *δ* ={0.00, 2.06, 2.57, 3.37, 4.18, 4.82,11.97, 16.15, 18.77, 21.12} ms; Δ = {0.00, 31.34, 31.85,32.65, 33.47, 34.10, 25.23, 29.41, 32.03, 34.38 }ms.

The scan was post-processed with routine pipelines[22], obtaining voxel-wise estimates of the vascular signal at *b* ≤100 s/mm^2^. Per-voxel maps of the top-ranking microvascular properties from the *in silico* study were obtained, using numerical models built on all synthetic networks. Standard IVIM vascular signal fraction and pseudo-diffusion coefficient (*f*_*V*_ and *D*^*^) were also obtained through segmented fitting[33, 22].

#### 2.2.2 Analysis: vascular metric characterisation

We computed mean and standard deviation of all metrics within regions-of-interest (ROIs) placed on: liver metastases (patient only); liver parenchyma; spleen.

An experienced radiologist (L.M.A.) identified the metastases.

## 3 Results

### 3.1 Vascular signal analysis *in silico*

#### 3.1.1 Microvascular networks

Supporting Information Fig. S1 visualises the 15 freely-available 2D networks from a previous study[22], and illustrates an example of 3D network obtained through the perturbation procedure.

#### 3.1.2 dMRI signal synthesis

Fig. 2 shows examples of dMRI signal decay as a function of *b* for three networks (panels (a) to (c)), as Δ varies. NC protocols are characterised by stronger signal decay than FC protocols. Moreover, the former show a strong dependence on Δ, unlike FC acquisitions. Non-monoexponential decay is seen for both, wide a range of signal attenuations.

**Figure 2.**
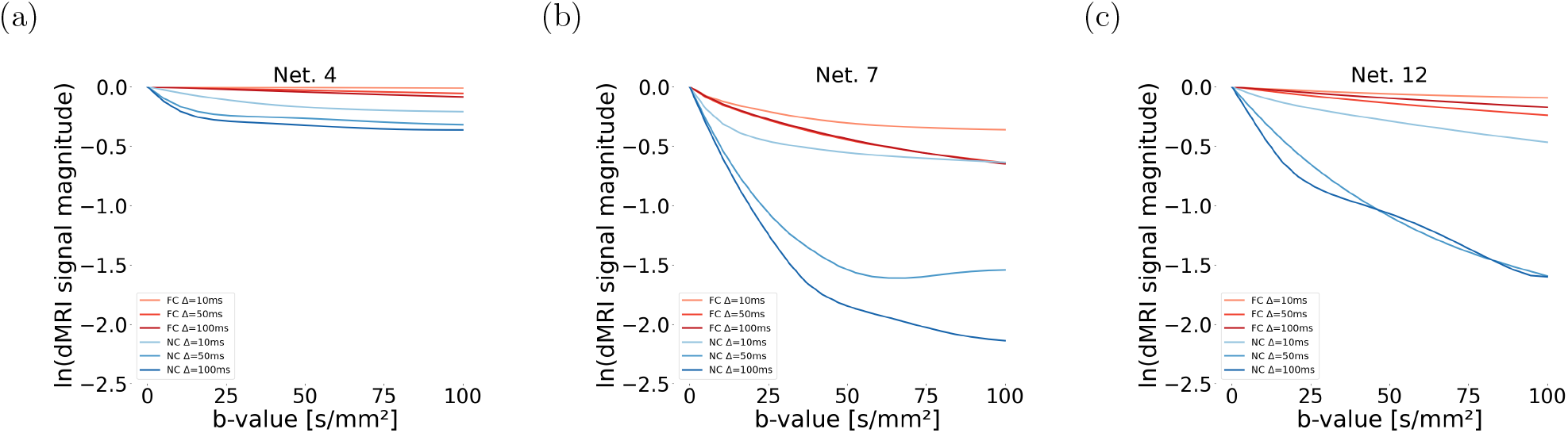
Illustration of the dMRI signal decay as a function of the b-value for three representative networks ((a): network 4; (b): network 7; (c): network 12). The figure reports decay for both NC and FC gradient waveforms, for different diffusion times. For the NC protocol: *δ* fixed to *δ* = 0.5 ms, Δ = {10, 50, 100} ms. For the FC protocol: *τ* fixed to = 1 ms, and Δ = {10, 50, 100 }ms. On the y-axis, we plot the logarithm of the synthetic vascular signal, to better highlight departures from the mono-exponential signal decay, which would be represented as a straight line. Non-monoexponential decay is seen for both protocols, wide a wide range of signal attenuations.

#### 3.1.3 Degrees of freedom estimation

Fig. 3 reports SVD for directionally-averaged signals for all protocols. The figure refers to the fixed diffusion time of *δ* = 6 ms and Δ = 30 ms for NC and richNC protocols; of *τ* = 10 ms and Δ = 30 ms for FC and richFC protocols. *N*_*p*_ = 2 independent SVs are detectable for SNR = 5 for all protocols, and *N*_*p*_ = 3 for SNR = 20.

**Figure 3.**
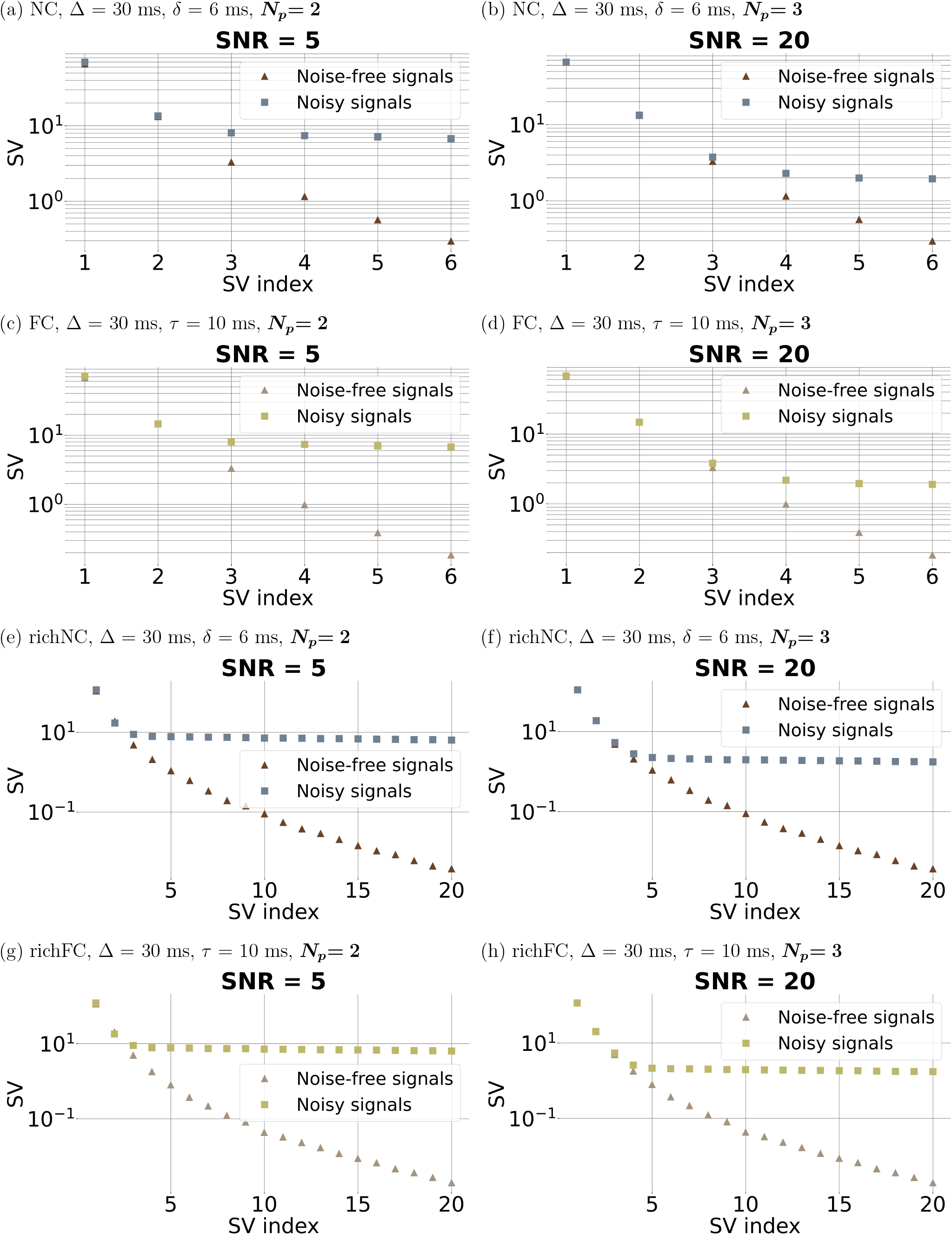
SVD for all non-compensated (NC) and flow-compensated (FC) protocols following directional averaging. Left, panels (a), (c), (e), (g): vascular dMRI sig_1_nal SNR of 5 at *b* = 0; right, panels (b), (d), (f), (h): vascular dMRI signal SNR of 20 at *b* = 0. From top to bottom: NC protocol ((a) and (b)); FC protocol ((c) and (d)); richNC protocol ((e) and (f)); richFC protocol ((g) and (h)). The figure refers to the fixed diffusion time of *δ* = 6 ms and Δ = 30 ms (protocols NC and richNC), and of *τ* = 10 ms and Δ = 30 ms (protocols FC and richFC).

Supporting Information Fig. S3 shows similar results for a different diffusion time (*δ* = 6 ms and Δ = 50 ms for the NC protocol; *τ* = 3 ms and Δ = 30 ms for the FC protocol). Results are in line to those of Fig. 3.

Fig. 4 shows SVs for the rich protocols when directionally averaging is not performed. SVD yields a considerably higher number of independent SVs: we detect *N*_*p*_ = 10 (richFC protocol) and *N*_*p*_ = 12 (richNC protocol) for SNR = 5, and *N*_*p*_ = 42 (richFC protocol) and *N*_*p*_ = 55 (richNC protocol) for SNR = 20.

**Figure 4.**
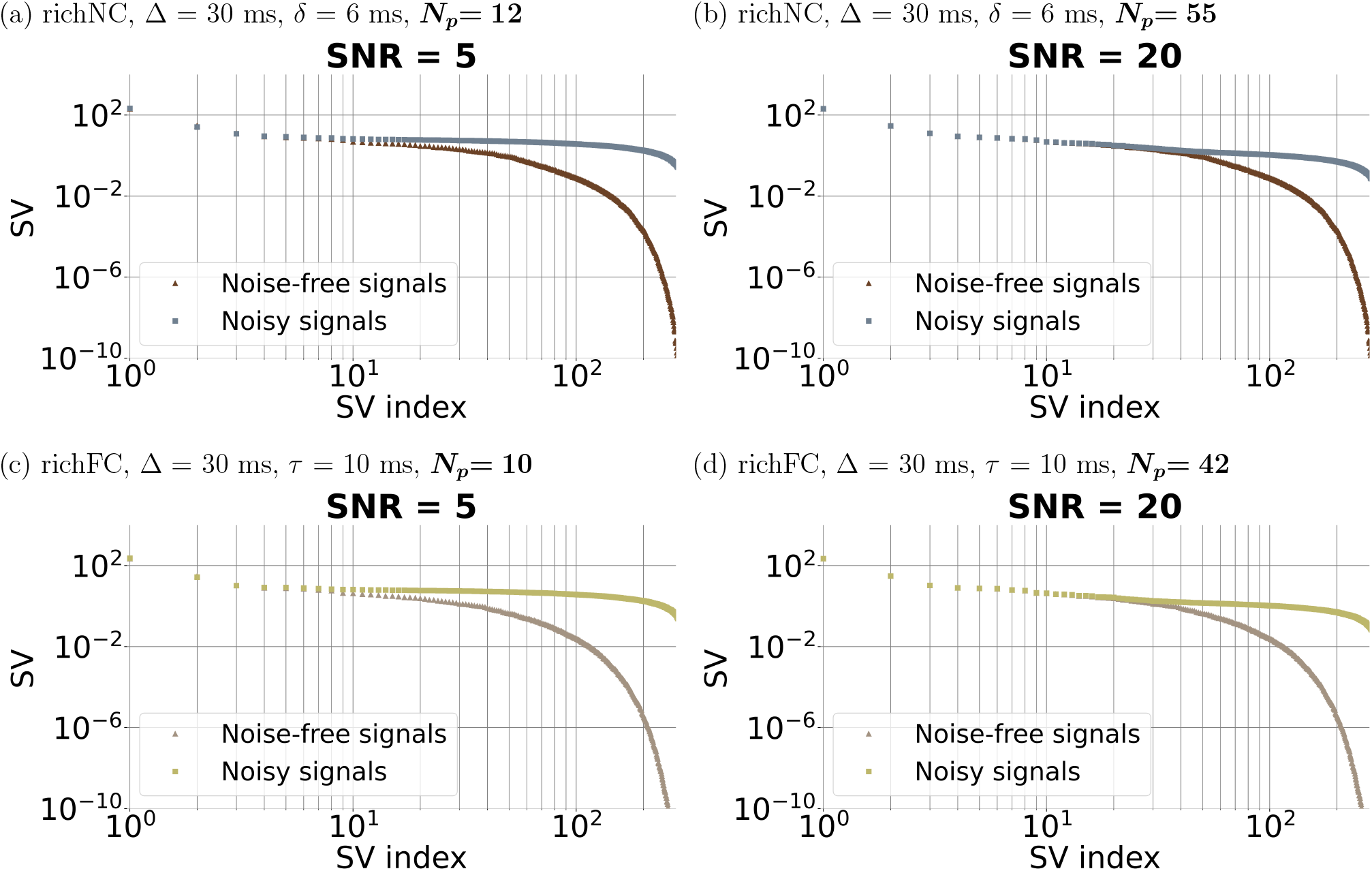
SVD for the rich non-compensated (richNC) and rich flow-compensated (richFC) protocols without directional averaging. Left, panels (a), (c): vascular dMRI signal SNR of 5 at *b* = 0; right, panels (b), (d): vascular dMRI signal SNR of 20 at *b* = 0. From top to bottom: richNC protocol ((a) and (b)); richFC protocol ((c) and (d)). The figure refers to the fixed diffusion time of *δ* = 6 ms and Δ = 30 ms (richNC), and of *τ* = 10 ms and Δ = 30 ms (richFC).

#### 3.1.4 Vascular property estimation and ranking

Fig. 5 shows scatter plots comparing ground truth against predicted vascular parameters for the NC protocol (Δ = 30 ms, *δ* = 6 ms, SNR of 5). The figure also reports Spearman’s correlation coefficients *r*_*s*_ and the BI for all metrics. Supporting Information Fig. S4 reports similar results for the FC protocol (Δ = 30 ms, *τ* = 10 ms). In both figures, the quality of the estimation varies greatly across parameters. Estimates of velocity and VFR distribution moments (e.g., *v*_*m*_ and *q*_*m*_) closely agree with ground truth values, unlike metrics related to the capillary geometry, which cannot be estimated (e.g., *r*_*s*_ of 0.681 for *q*_*m*_ against 0.025 for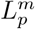). Strong correlations with ground truth and limited bias (BI *<* 8 %) are achieved for *ANB* in both protocols. Higher values of *r*_*s*_ are observed for the FC than the NC protocol (e.g., *r*_*s*_ of 0.453, 0.681 and 0.653 for *v*_*m*_, *q*_*m*_ and *ANB* for NC, while of 0.496, 0.733, 0.668 for the FC).

**Figure 5.**
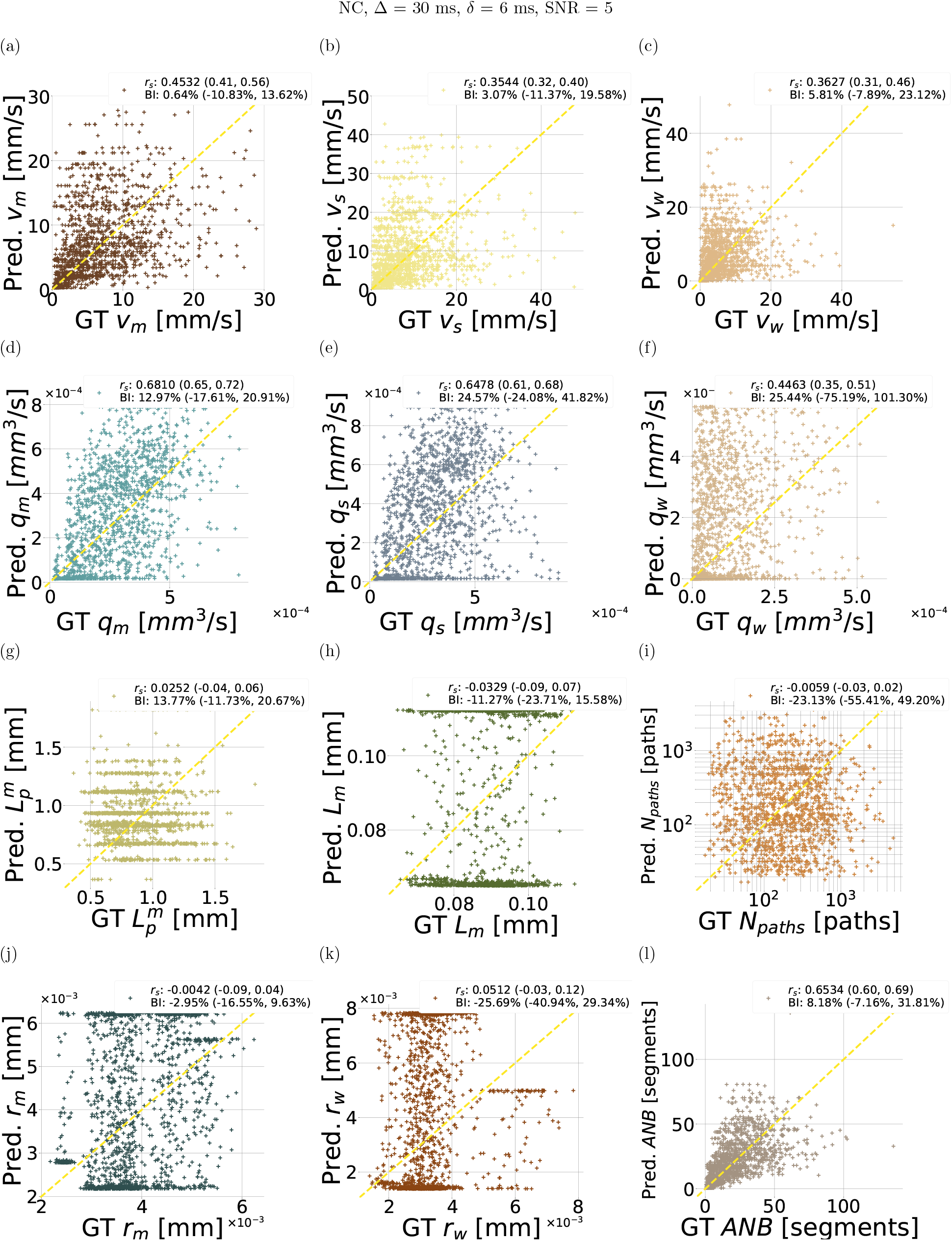
Scatter plots of estimated vascular parameters against ground truth values from the leave-one-out fitting procedure implemented *in silico*. The figure refers to the NC protocol, with Δ = 30 ms and *δ* = 6 ms, SNR = 5. From top to bottom: first row, mean velocity *v*_*m*_ in (a), standard deviation of velocity *v*_*s*_ in (b), path-weighted mean velocity *v*_*w*_ in (c); second row,_1_mean volumetric flow rate (VFR) *q*_*m*_ in (d), standard deviation of VFR *q*_*s*_ in (e), path-weighted mean VFR *q*_*w*_ in (f); third row, mean input/output path length 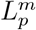 in (g), mean capillary segment length *L*_*m*_ in (h), number if input/output paths *N*_*paths*_ in (i); fourth row, mean capillary radius *r*_*m*_ in (j), path-weighted mean capillary radius *r*_*w*_ in (k), and apparent network branching *ANB* in (l). For each metric, the overall Spearman’s correlation coefficient *r*_*s*_ and Bias Index (BI) are reported, with the range of *r*_*s*_ and BI values obtained across leave-one-out folds. “GT” and “Pred.” respectively indicate ground truth and predicted metric values.

Supporting Information Fig. S5 and Fig. S6 show scatter plots for SNR = 20. For *v*_*m*_, an increase in SNR is associated to a clear increase in the correlation *r*_*s*_ (from 0.453 to 0.573), and a decrease in BI range across folds. *v*_*s*_ and *v*_*w*_ show similar trends. VFR parameters (*q*_*m*_, *q*_*s*_, *q*_*w*_) generally maintain stronger correlations across both SNR conditions, which also increase as SNR increases (e.g., *r*_*s*_ = 0.681, SNR = 5 and *r*_*s*_ = 0.751, SNR = 20 for *q*_*m*_), with minor changes in the BI. Geometry parameters 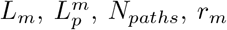 and *r*_*w*_ show weak correlation between estimated and ground truth values even for SNR = 20. *ANB* estimation greatly benefits from increased SNR: *r*_*s*_ grows from 0.653 (SNR of 5) to 0.715 (SNR of 20), and the BI range across folds is narrowed ((-7.61%,31.81%) for SNR = 5 to (-2.83%,22.38%) for SNR = 20).

Supporting Information Fig. S7 and S8 shows estimation results for a second diffusion time (longer, for the NC protocol: Δ = 50 ms, *δ* = 6 ms; shorter, for the FC protocol: Δ = 30 ms, *τ* = 3 ms; SNR = 20). While results are consistent with all the above, we also note that in general estimation performances improve as the diffusion time increases (e.g., compare: NC protocol, *r*_*s*_ for *v*_*m*_ of 0.573 for Δ = 30 ms and of 0.623 for Δ = 50 ms; FC protocol, *r*_*s*_ for *v*_*m*_ of 0.438 for *τ* = 3 ms and of 0.676 for *τ* = 10 ms).

Fig. 6 and Supporting Information Fig. S9 show rankings of metrics obtained based on the Spearman’s correlation *r*_*s*_. Each figure reports rankings for NC, FC and hybrid protocols (Fig. 6: NC Δ = 30 ms, *δ* = 6 ms; FC Δ = 30 ms, *τ* = 10 ms; Supporting Information Fig. S9: NC Δ = 50 ms, *δ* = 6 ms; FC Δ = 30 ms, *τ* = 3 ms). Similar rankings are also reported for the rich protocols in Supporting Information Fig. S10. In all cases, *q*_*m*_, *q*_*s*_ and *ANB* are the top-ranking metrics, albeit with slightly different orders depending on the protocol and SNR. The adoption of a hybrid FC/NC acquisition strategy positively impacts on the ranking figures, especially at the low SNR level of 5, where the estimation of *q*_*m*_, *ANB* and *q*_*s*_ outperforms FC and NC protocols.

**Figure 6.**
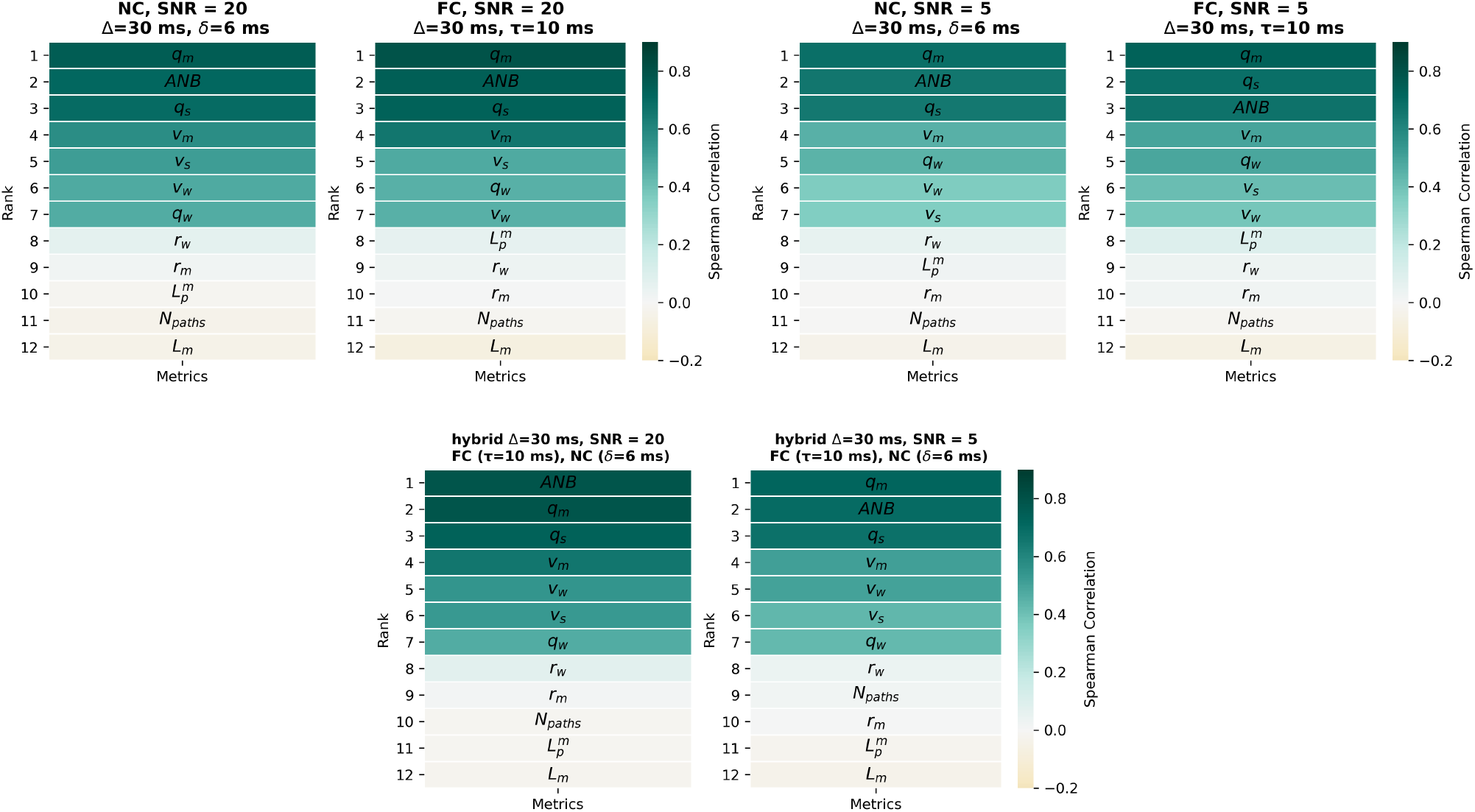
Ranking of microvascular metrics based on the goodness of the estimation, i.e., based on sorting the Spearman’s correlation *r*_*s*_ coefficients of all microvascular properties. For each property, *r*_*s*_ was computed by relating ground truth and estimated metric values. Ranking was obtained for the NC and FC protocols for SNR = 20 and SNR = 5, and for a hybrid protocol alternating measurements from the NC and the FC gradient waveforms. Top row, from left to right: ranking for the NC protocol (SNR = 20), FC protocol (SNR = 20), NC protocol (SNR = 5), FC protocol (SNR = 5). Bottom row, from left to right: ranking for the hybrid protocol at SNR = 20 and SNR = 5. The figure refers to the following diffusion time: Δ = 30 ms, *δ* = 6 ms for the NC protocol; Δ = 30 ms, *τ* = 10 ms for the FC protocol.

Supporting Information Fig. S11 shows a final example in which *q*_*m*_ and *ANB* are estimated jointly, with performances comparable to those obtained when the metrics are estimated individually (all cases above).

*Given these consistent trends, in the following material we will focus on the estimation of VFR distribution moments and of ANB. When considering two tissue parameters, we will focus on q*_*m*_ *(first VFR moment) and on ANB*.

#### 3.1.5 Relationship between vascular properties and vascular signal features

Fig. 7 shows the dependence of metrics *q*_*m*_ and *ANB* on salient vascular dMRI signal cumulants *D*^*^ and *K*^*^ (apparent vascular diffusion and kurtosis coefficients).

**Figure 7.**
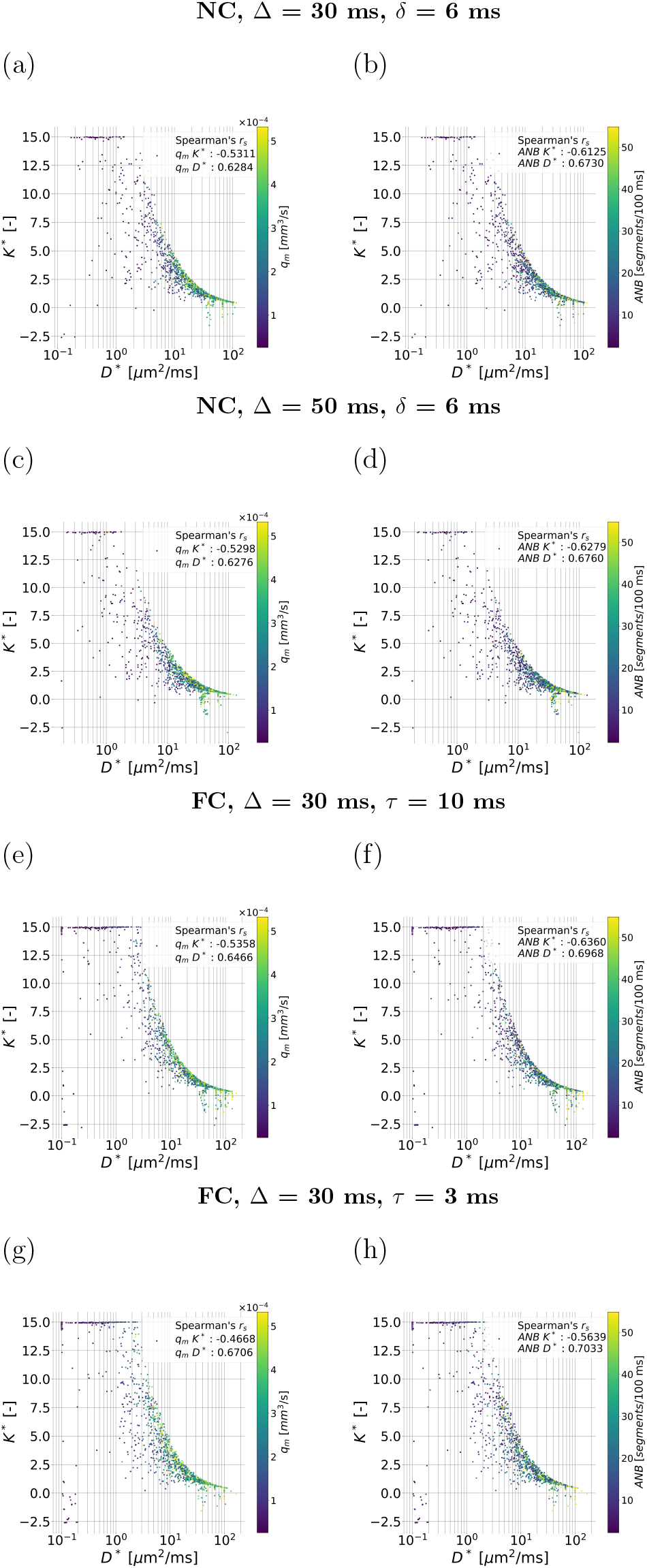
Scatter plots visualising the relationship between the vascular dMR signal cumulants and the topranking metrics selected from the *in silico* study. The figure scatters the apparent vascular diffusion and kurtosis coefficients (*D*^*^ and *K*^*^) against each other, colouring the points according to the mean VFR *q*_*m*_ and the apparent network branching *ANB*. Panels (a) and (b), top row: NC protocol, Δ = 30 ms, *δ* = 6 ms (*q*_*m*_ = *f* (*D*^*^, *K*^*^) in (a); *ANB* = *f* (*D*^*^, *K*^*^) in (b)). Panels (c) and (d), second row: as (a) and (b), but for Δ = 50 ms, *δ* = 6 ms. Panels (e) and (f), third row: FC protocol, Δ = 30 ms, *τ* = 10 ms (*q*_*m*_ = *f* (*D*^*^, *K*^*^) in (e); *ANB* = *f* (*D*^*^, *K*^*^) in (f). Panels (g) and (h), fourth row: as (e) and (f), but for Δ = 50 ms, *τ* = 3 ms. Spearman’s correlation coefficients between *q*_*m*_ and *D*^*^ and *K*^*^, and between *ANB* and *D*^*^ and *K*^*^, are also reported.

For both NC and FC protocols and for multiple dif-fusion times, there is a moderate-to-strong association between *D*^*^, *K*^*^ and either of *q*_*m*_, *ANB*. The correlation strength is similar for both diffusion times for a given protocol, and, in general, between protocols (e.g., NC protocol: *D*^*^/*q*_*m*_ Spearman’s correlation *r*_*s*_ of 0.628 for Δ = 50 ms; FC protocol: *D*^*^/*q*_*m*_ Spearman’s correlation *r*_*s*_ of 0.671 for *τ* = 3 ms). *D*^*^ increases for increasing diffusion time.

Supporting Information Fig. S12 includes similar plots for the rich protocols. The same trends are seen. Additionally, the figure highlights that while *q*_*m*_ and *ANB* are positively correlated (*r*_*s*_ of 0.718), a range of *ANB* values can be observed for any given *q*_*m*_.

### 3.2 Vascular signal analysis *in vivo*

#### 3.2.1 Vascular metric characterisation

Fig. 8 shows MRI data from the metastatic CRC patient at baseline, and after 3 months of targeted therapy against liver metastases. The figure reveals the location of the metastasis within the liver parenchyma, and demonstrates that these are replaced by healed parenchymal tissue after treatment. The figure also depicts *q*_*m*_ and *ANB* maps. At baseline, on visual inspection both metrics appear lower in the metastases than in the liver. At follow-up, both *q*_*m*_ and *ANB* increases, recovering values similar to those of the liver parenchyma.

**Figure 8.**
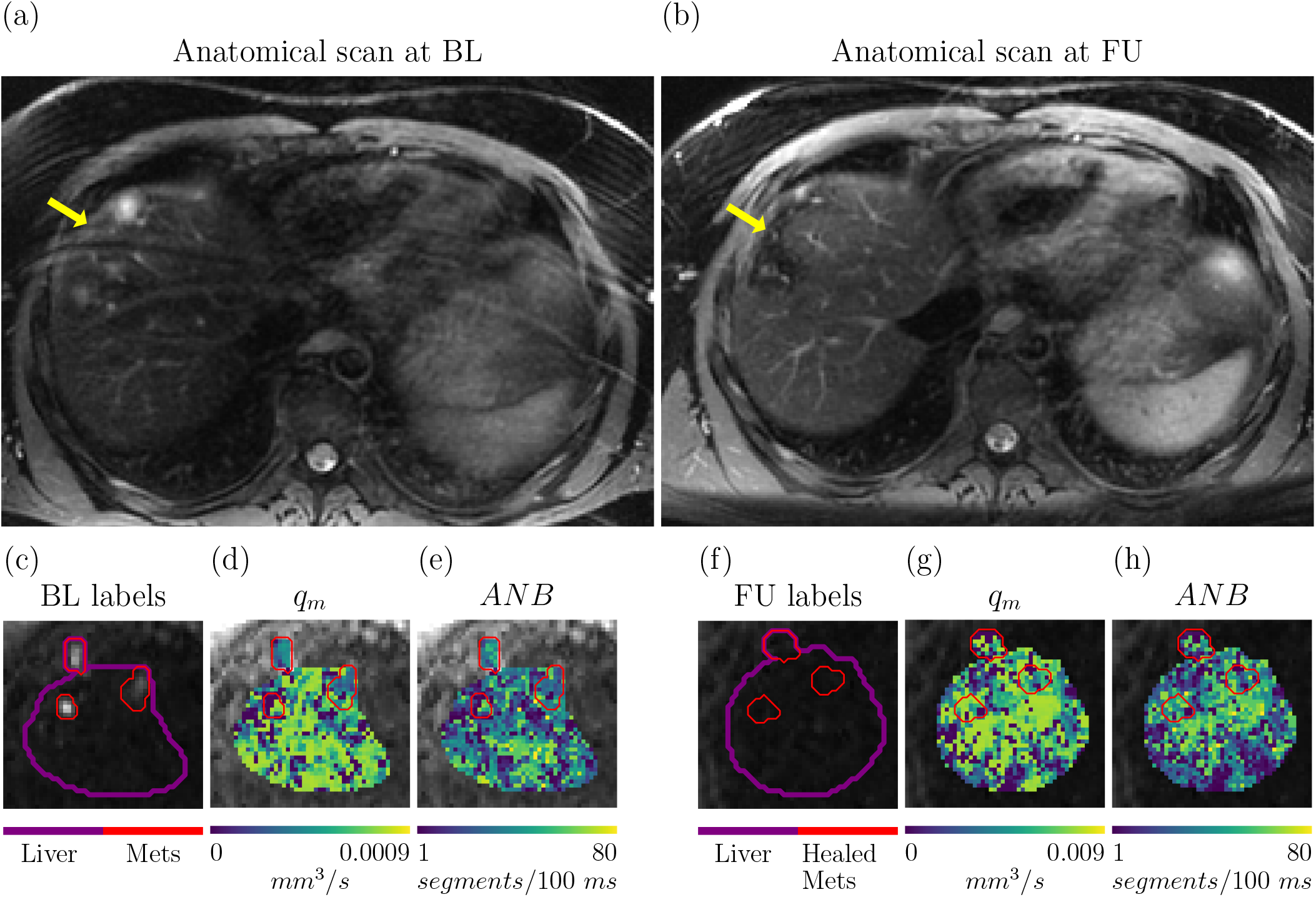
Microvascular parameter estimation in a CRC patient *in vivo*. Top: structural, anatomical highresolution T2-weighted scan of the liver obtained at baseline (BL) and at follow-up (FU), after 3 months of targeted therapy for metastatic CRC. (a): BL scan, with arrow illustrating the position of two metastases. (b): FU scan, highlighting the effect of treatment, with healed liver parenchyma replacing the metastases. Bottom: microvascular maps *q*_*m*_ (mean VFR) and *ANB* (apparent network branching) at the two time points. From left to right, (c): high b-value image highlighting the position of 3 CRC metastases within the liver at BL; (d) *q*_*m*_ map at BL; (e): *ANB* map at BL; (f): high b-value image at FU, in the same liver location featuring metastases at BL, now featuring healed tissue; (g) *q*_*m*_ map at FU; (h): *ANB* map at FU.

Table 1 reports mean and standard deviation of *q*_*m*_, *ANB* and of IVIM *f*_*V*_ and *D*^*^ in multiple ROIs. Quantitative trends confirm what was observed on visual inspection. The liver features higher vascularisation than both spleen and metastases (higher *f*_*V*_, *D*^*^, *q*_*m*_ and *ANB*). After treatment, all vascular metrics in the healed metastases increase, and hence feature values that match closely those of the liver parenchyma ROI.

**Table 1:** Mean and standard deviation of metrics from simulation-informed and IVIM parameter fitting in different ROIs. The table report metrics for the colorectal cancer (CRC) and for the healthy volunteer. For the CRC case, results from both baseline (BL) scan and follow-up (FU, consisting in 3 months of targeted therapy against CRC liver metastases) are included.

| ID | ROI | $q_m$ [$10^3$ mm<sup>3</sup>/s] | ANB [segments/100 ms] | $f_v$ | $D^*$ [$\mu\text{m}^2/\text{ms}$] |
| --- | --- | --- | --- | --- | --- |
| Patient at BL |  |  |  |  |  |
|  | Metastases | 0.279 (0.170) | 23.9 (15.3) | 0.230 (0.217) | 8.70 (16.5) |
|  | Spleen | 0.305 (0.283) | 28.7 (20.6) | 0.185 (0.213) | 15.1 (38.6) |
|  | Liver | 0.539 (0.270) | 38.3 (22.5) | 0.598 (0.298) | 18.7 (35.7) |
| Patient at FU |  |  |  |  |  |
|  | Healed metatstases | 0.571 (0.229) | 41.7 (19.7) | 0.384 (0.335) | 20.6 (38.8) |
|  | Spleen | 0.446 (0.290) | 37.7 (20.9) | 0.131 (0.178) | 24.8 (48.9) |
|  | Liver | 0.565 (0.244) | 38.8 (21.0) | 0.455 (0.316) | 23.0 (47.5) |
| Volunteer |  |  |  |  |  |
|  | Spleen | 0.439 (0.252) | 39.9 (18.7) | 0.123 (0.124) | 23.7 (43.3) |
|  | Liver | 0.542 (0.215) | 49.1 (16.6) | 0.359 (0.227) | 45.9 (62.9) |

Table 1 also lists statistics in the healthy healthy volunteer’s spleen and liver ROI, which are in line with values seen in the patient (maps in Supporting Information Fig. S13).

## 4 Discussion

### 4.1 Summary

In this article, we used simulations of blood flow in synthetic capillary networks to study microvascular parameter mapping. We simulated a variety of dMRI protocols, which included compact or rich b-value samplings, multiple diffusion times and gradient directions, with NC and FC encodings. The synthetic signals reveal a variety of contrasts across networks, with complex signal features. SVD suggests that 2 to 3 independent degrees of freedom are encoded in the directionally-average signal in presence of noise, a number that goes up to 10 to 50 without directional averaging.

Numerical models developed from the synthetic signals suggest that the VFR distribution (e.g., mean VFR *q*_*m*_) and the *ANB*, characterising network complexity, are the parameters that can be more robustly inferred from noisy signal measurements. Adequate sampling of the vascular signal appears key for their estimation, since both pseudo-diffusion and kurtosis coefficients (*D*^*^ and *K*^*^) are modulated by *q*_*m*_ and *ANB*. We also tested whether *q*_*m*_ and *ANB* could be estimated *in vivo* on a healthy volunteer and a CRC patient with liver metastases. We studied *q*_*m*_ and *ANB* within metastasis, liver, and spleen ROIs, alongside classical IVIM *f*_*V*_ and *D*^*^. The metrics reveal vascularisation differences between the metastases and the liver, with lower *q*_*m*_, *ANB, f*_*V*_ and *D*^*^ in the former. These differences are reversed after 3 months of targeted therapy, showing potential for treatment monitoring.

### In silico study

We used freely available vascular networks from a previous study[22] to generate rich dictionaries of synthetic vascular dMRI signals, with coupled microvascu-lar properties, for a variety of acquisition settings. We processed the signals (i) to estimate the number of independent microvascular degrees of freedom that can be potentially recovered at realistic SNR; (ii) to discover which microvascular properties can be inferred in practice; (iii) to characterise salient features of the vascular signal.

The estimation of the degrees of freedom, based on signal matrix SVD, demonstrates that at realistic SNR (e.g., between 5 to an optimistic upper bound of 20[34] on the vascular signal), 2-3 independent components can be captured with directionally-averaged signals. This number increases to 10 to 50, in the best SNR case of 20, without directional averaging. These findings suggest that around 3 fully independent microvascular parameters can be practically estimated from clinical dMRI acquisitions, which typically use directionally-averaged trace imaging. When richer sets of gradient directions available instead, a higher number of significant SVs may be detected. Noting that the vascular pseudo-diffusion tensor (first order cumulant) contains 6 independent parameters[35], this implies that higherorder directional signal features (e.g., vascular kurtosis tensors[36, 37] could be exploited for the estimation of tensorial extensions of the scalar metrics considered here.

Afterwards, we investigated which microvascular properties could be estimated in practice from noisy signal measurements. Maximum-likelihood fitting of numerical signal models suggests that for a variety of acquisition settings, the moments of the VFR distribution (*q*_*m*_ and *q*_*s*_) and the *ANB* are the metrics that can be most robustly estimated. *For this reason q*_*m*_ *and ANB are the properties we recommend focussing estimation efforts on in non-invasive microvascular mapping, when two metrics are of interest*. Interestingly, these results also suggest that the VFR distribution can be better recovered than the blood velocity distribution. This results, heretofore undescribed, shows the potential of physics-informed simulations to guide the design of new MRI biomarkers.

Conversely, metrics related to the network geometry, as the mean capillary radius/length, cannot be recovered even at high SNR. This likely results from the fact that the VFR *q* and the velocity **v** are directly related the spin trajectories **p** encoded in the spin phase, 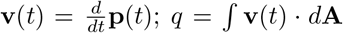 where *d***A** is the elementary area element). Conversely, indices of capillary geometry alone (e.g., radius or length) are not sufficient to determine the overall flow resistance offered by a network, and hence the corresponding VFR field and the spin trajectories, unless information capillary arrangement (e.g., serial vs parallel [22]) is known.

The comparison between estimated and ground truth vascular parameters highlights other notable facts. For example, the choice of the diffusion time impacts the quality of parameter estimation, with longer diffusion times yielding better performances for both NC and FC acquisitions. Moreover, hybrid acquisitions that combine both NC and FC measurements outperform FC and NC protocols. This is in line with known literature[16, 38, 39], since using both type of acquisition increases the contrast across DW measurements.

We also characterised salient features of the vascular dMRI signal across multiple protocols. This analysis reveals a clear non-mono-exponential behaviour of the signal as a function of *b* (non-zero kurtosis). It also highlights diffusion-time dependence, in agreement with previous studies[22], and shows that both vascular pseudo-diffusion and kurtosis coefficients (*D*^*^ and *K*^*^) are sensitive to *q*_*m*_ and *ANB*. This result stresses the importance of adequate b-value sampling design for robust microvascular parameter mapping, in order to capture not only the slope of the (log)-signal decay (*D*^*^), but also changes in signal curvature (*K*^*^).

Lastly, further analyses show that *q*_*m*_ and *ANB* are positively correlated between each other. This implies that additional vascular parameters could be potentially be inferred jointly with *q*_*m*_ and *ANB* (recall that to 2-3 *fully independent* degrees of freedom appear to be detectable). Nevertheless, we point out that the joint estimation of *ANB* and *q*_*m*_ is not completely redundant, since each of these metrics adds some information to the other: note that a range of *ANB* values can be observed for any *q*_*m*_ in Supporting Information Fig. S12.

### 4.3 In vivo study

We also tested whether the most promising vascular metrics from the *in silico* analysis — i.e., *q*_*m*_ and *ANB* —, could be measured on human volunteers *in vivo*, on clinical MRI hardware. With this goal in mind, we acquired dMRI data on a healthy volunteer and on a CRC patient, treated with targeted therapy against liver metastases. Scans were acquired at 3T, with the patient scanned before treatment administration, and after 3 months of therapy.

Results suggest the feasibility of estimating these metrics *in vivo*. While *ANB* mapping was previously demonstrated in our recent study[22], here we provide further evidence towards its robustness across protocols. Regarding VFR, to our knowledge this is the first time that VFR moments have been mapped *in vivo* with dMRI. Both *ANB* and VFR show trends that are plausible, and in line with well-established IVIM indices. Importantly, our new metrics capture temporal changes potentially associated treatment effects, with healed tissue recovering flow properties of the liver parenchyma, highlighting the potential of physics-informed microvascular dMRI modelling.

### 4.4 Methodological considerations and limitations

Several aspects of this study deserve further considerations. Firstly, our simulations are based on a limited sample of small networks[22]. We acknowledge that further confirmation is required from the simulation of larger and more complex networks, obtained, for example, through generative approaches[40] or from 3D histology[41, 42].

Secondly, in our *in silico* experiments we analysed signals with different b-value samplings, and performed metric ranking on multiple diffusion times. This was done to ensure that results where not specific to a single acquisition, striving for generalisability. However, we acknowledge that further work is required: (i) to fully characterise the impact of Δ, *δ, G*, and *τ* on vascular metric estimation; (ii) to design acquisition protocols that maximise the signal sensitivity to a target metric of interest, e.g., through Cramer-Rao Lower Bound optimisation[43]; (iii) to assess the estimation of tensorial extensions of the vascular metrics considered here.

Regarding the dMRI protocols, our simulations highlighted the benefits of combining FC and NC measurements. The use of FC waveforms may be especially beneficial *in vivo*, since these compensate for some of the unavoidable patient’s bulk motion [38]. However, it should be remembered the FC waveforms may suffer from reduced diffusion-weighting efficiency, and decreases in SNR[44].

Lastly, we acknowledge that our pilot cohort is small, and that validation in larger patient data bases is required to enable clinical translation.

### 4.5 Conclusions

Physics-based simulations of the vascular dMRI signal inform the design of innovative microperfusion biomarkers for *in vivo* imaging. The estimation of two to three fully independent vascular parameters from IVIM-like acquisitions appears feasible *in vivo*, with VFR distribution moments and indices of vascular network branching being the most promising metrics. These may enable the non-invasive assessment of body tumour characteristics, and play a key role in precision oncology.

## Data Availability

The simulated data and the code will be made available at the following link: https://github.com/radiomicsgroup/SpinFlowSim.
The human data cannot be made freely available due to ethical issues. Authors interested in accessing the data can contact the corresponding author(s) to establish appropriate, institutional data transfer and sharing agreements.

## Acknowledgments

VHIO would like to acknowledge: the State Agency for Research (Agencia Estatal de Investigación) for the financial support as a Center of Excellence Severo Ochoa (CEX2020-001024S/AEI/10.13039/501100011033), the Cellex Foundation for providing research facilities and equipment; the CERCA Programme from the Generalitat de Catalunya for their support on this research. This study has been co-funded by the European Regional Development Fund/European Social Fund ‘A way to make Europe’ (to R.P.L.), and by the Comprehensive Program of Cancer Immunotherapy and Immunology (CAIMI), funded by the Banco Bilbao Vizcaya Argentaria Foundation Foundation (FBBVA, grant 89/2017). R.P.L. is supported by the “la Caixa” Foundation CaixaResearch Advanced Oncology Research Program, the Prostate Cancer Foundation (18YOUN19), a CRIS Foundation Talent Award (TALENT19-05), the FERO Foundation through the XVIII Fero Fellowship for Oncological Research, the Instituto de Salud Carlos III-Investigación en Salud (PI18/01395 and PI21/01019), the Asociación Española Contra el Cancer (AECC) (PRYCO211023SERR) and the Generalitat de Catalunya Agency for Management of University and Research Grants of Catalonia (AGAUR) (2023PROD00178, SGR-Cat2021). The project that gave rise to these results received the support of a fellowship from “la Caixa” Foundation (ID 100010434). The fellowship code is “LCF/BQ/PR22/11920010” (funding F.G., A.V., and A.G.). M.P. is supported by the UKRI Future Leaders Fellowship MR/T020296/2.

A.G. is supported by a Severo Ochoa PhD fellowship (PRE2022-102586; Plan Estatal de Investigación Científica, Técnica y de Innovación 2022, Agencia Estatal de Investigación (AEI)). The authors are thankful to the ASCIRES CETIR clinical team for their assistance with MRI acquisitions, and to the GE clinical scientists for their support with diffusion sequence characterisation. We also warmly thank healthy volunteers, patients and their families, for the time and effort they commit to research.

## Supporting Information

**Supporting Information Figure S1:**
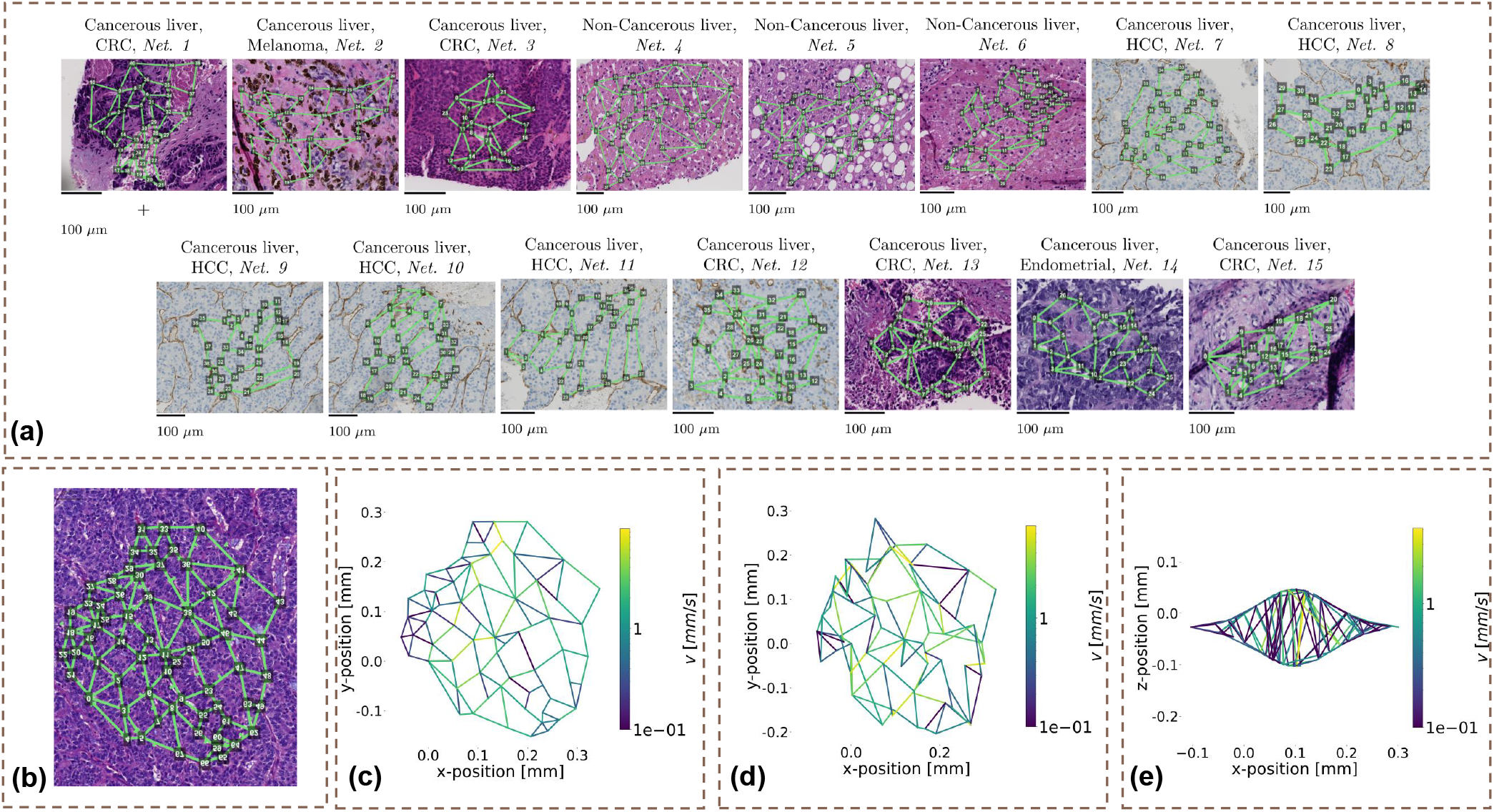
Illustration of the vascular networks used in our article. (a): the 15 2D freely available networks from a previous study[22] processed for our experiments *in silico*. (b)-(d): illustration of the perturbation procedure followed to generate a rich set of 1500 tridimensional and unique networks. (b): 2D network; (c): 2D network with resolved velocity field; (d): the same network after perturbation of radii and node positions; (e): final 3D shape of the network. Panel (a) has been generated with permission by adapting Figure 2 of Voronova et al[22], Medical Image Analysis 2025, 102: 103531, doi: 10.1016/j.media.2025.103531. Journal: Medical Image Analysis, Elsevier (https://www.sciencedirect.com/journal/medical-image-analysis). License number 6078220596843 obtained on July 29th 2025 through CCC RightsLink.

**Supporting Information Figure S2:**
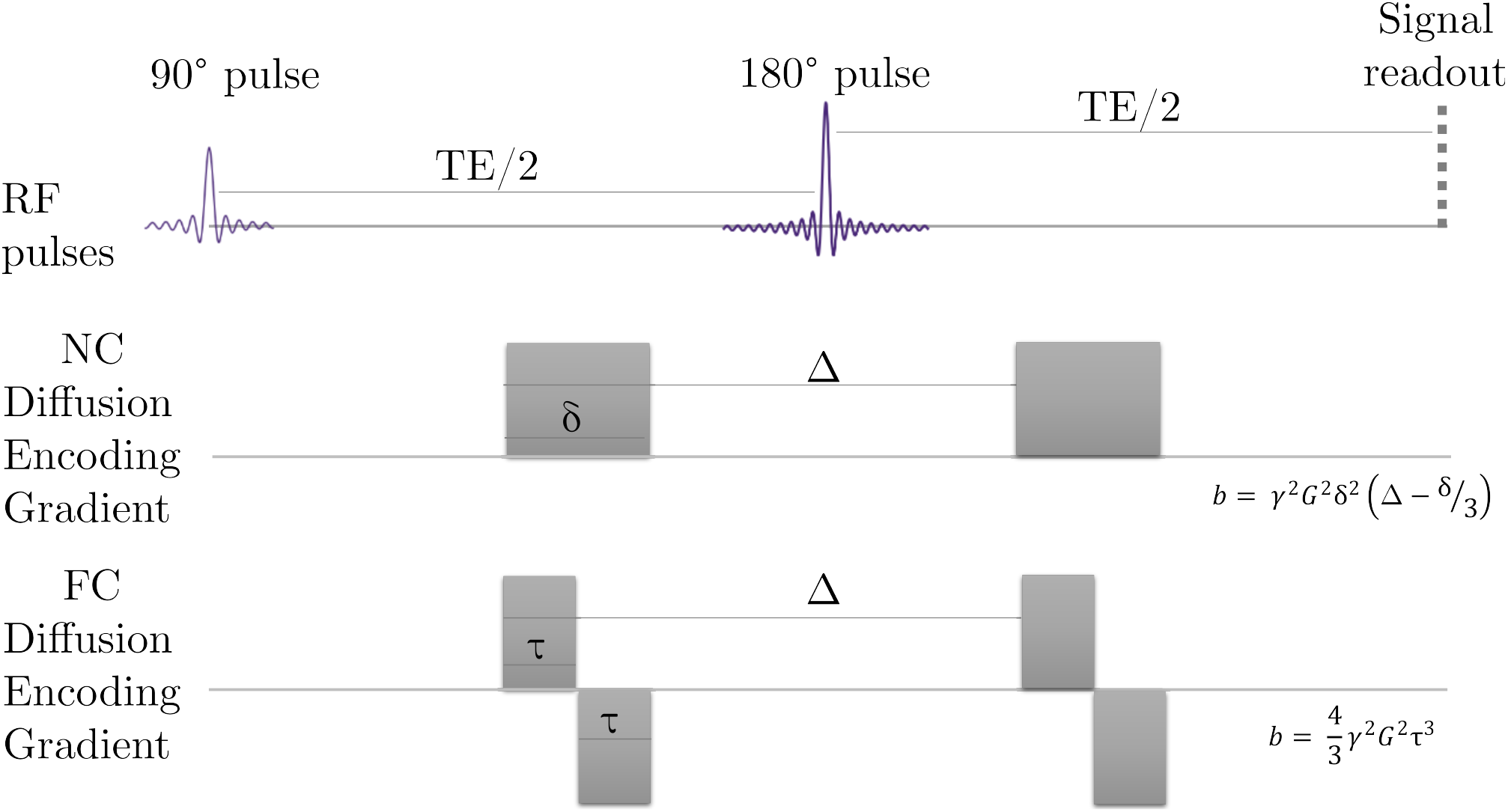
Illustration of the NC and FC gradient waveforms used in this study, with corresponding b-value expressions.

**Supporting Information Figure S3:**
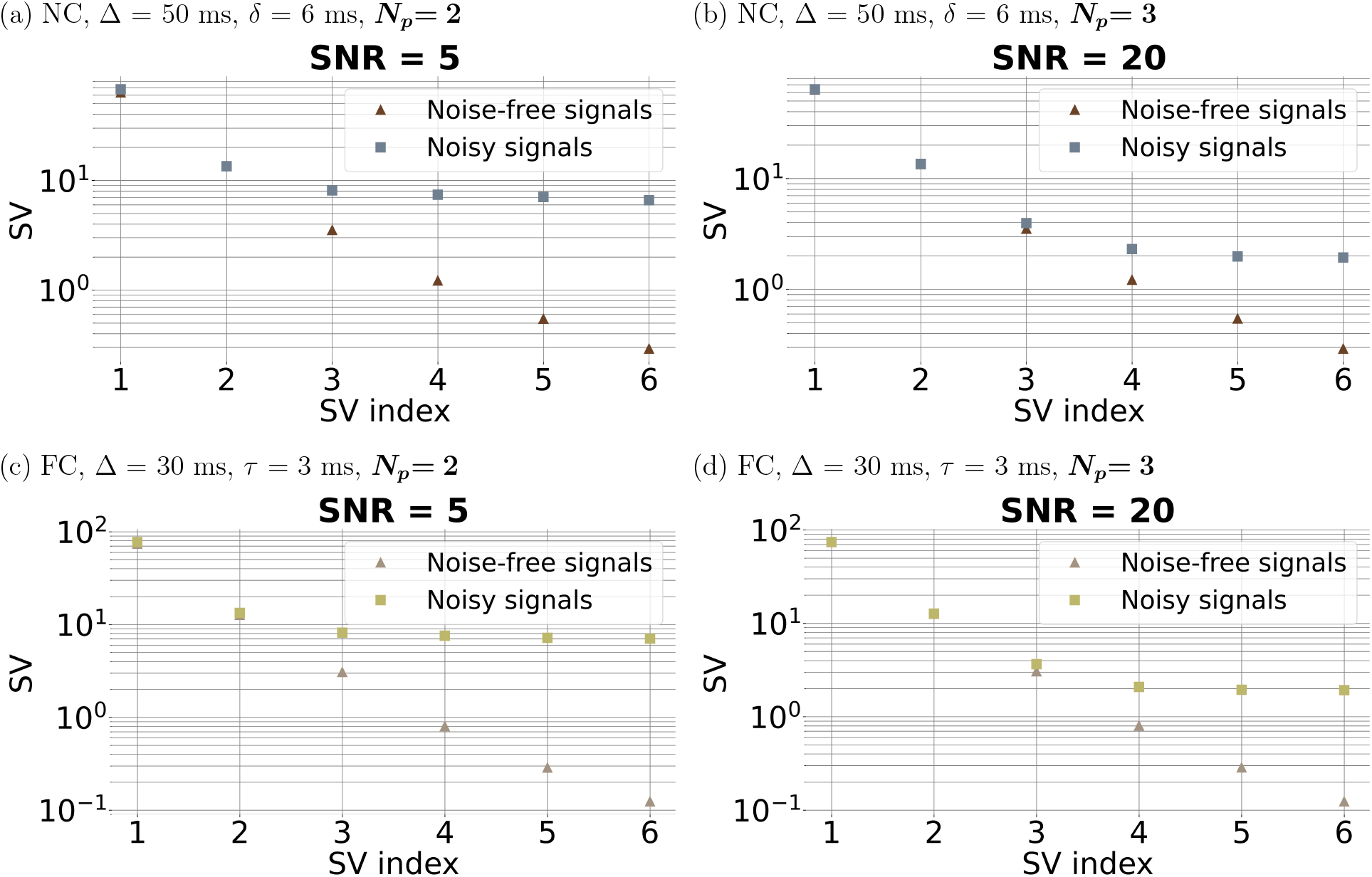
SVD for the non-compensated (NC) and flow-compensated (FC) protocols following directional averaging for a second diffusion time. Left, panels (a), (c): vascular dMRI signal SNR of 5 at *b* = 0; right, panels (b), (d): vascular dMRI signal SNR of 20 at *b* = 0. From top to bottom: NC protocol ((a) and (b)); FC protocol ((c) and (d)). The figure refers to the fixed diffusion time of *δ* = 6 ms and Δ = 50 ms (NC protocol), and of *τ* = 3 ms and Δ = 30 ms (FC protocol).

**Supporting Information Figure S4:**
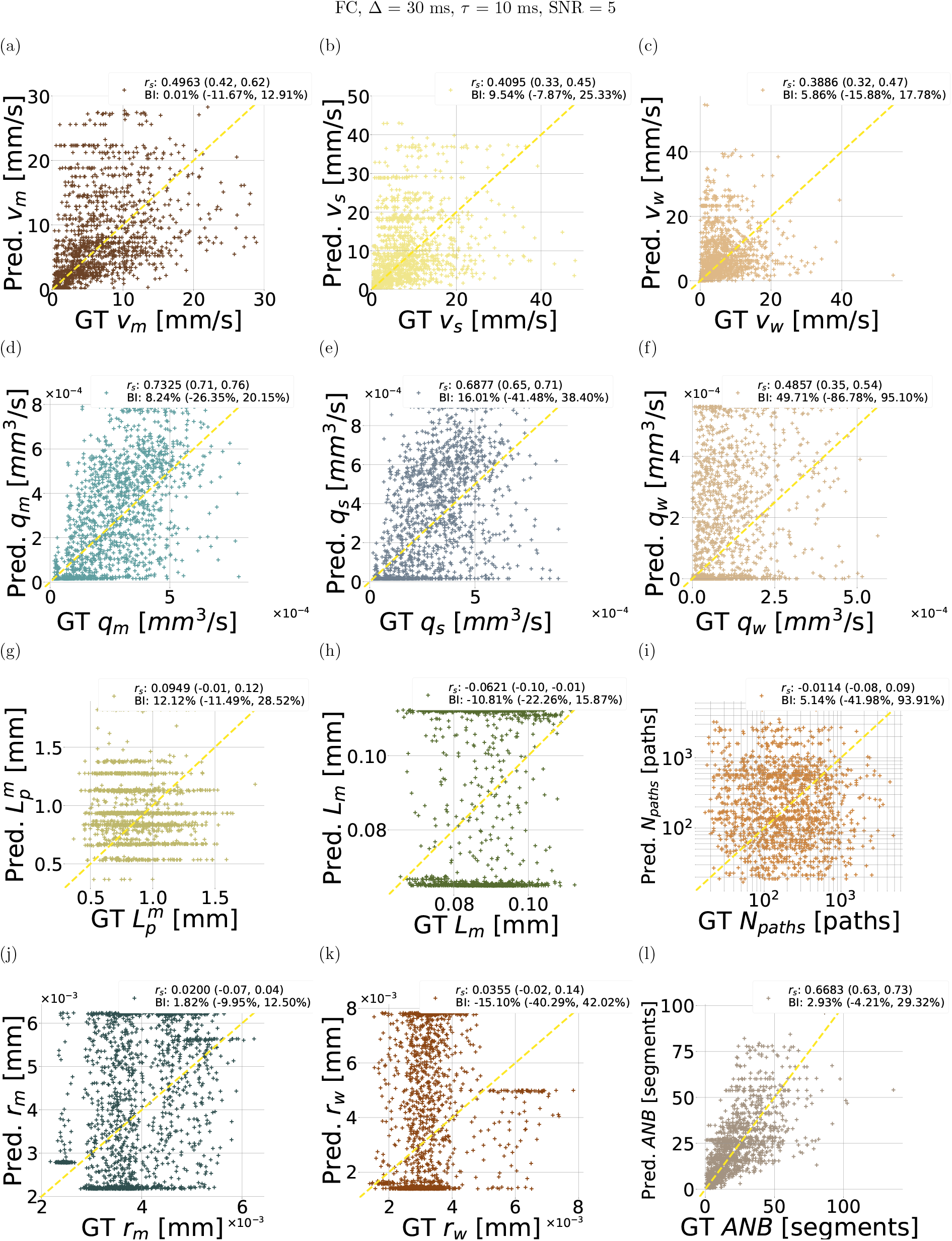
Scatter plots of estimated vascular parameters against ground truth values from the leave-one-out fitting procedure implemented *in silico*. The figure refers to the FC protocol, with Δ = 30 ms and *τ* = 10 ms, SNR = 5. From top to bottom: first row, mean velocity *v*_*m*_ in (a), standard deviation of velocity *v*_*s*_ in (b), path-weighted mean vel^1^ocity *v*_*w*_ in (c); second row, mean volumetric flow rate (VFR) *q*_*m*_ in (d), standard deviation of VFR *q*_*s*_ in (e), path-weighted mean VFR *q*_*w*_ in (f); third row, mean input/output path length 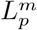 in (g), mean capillary segment length *L*_*m*_ in (h), number if input/output paths *N*_*paths*_ in (i); fourth row, mean capillary radius *r*_*m*_ in (j), path-weighted mean capillary radius *r*_*w*_ in (k), and apparent network branching *ANB* in (l). For each metric, the overall Spearman’s correlation coefficient *r*_*s*_ and Bias Index (BI) are reported, with the range of *r*_*s*_ and BI values obtained across leave-one-out folds.. “GT” and “Pred.” respectively indicate ground truth and predicted metric values.

**Supporting Information Figure S5:**
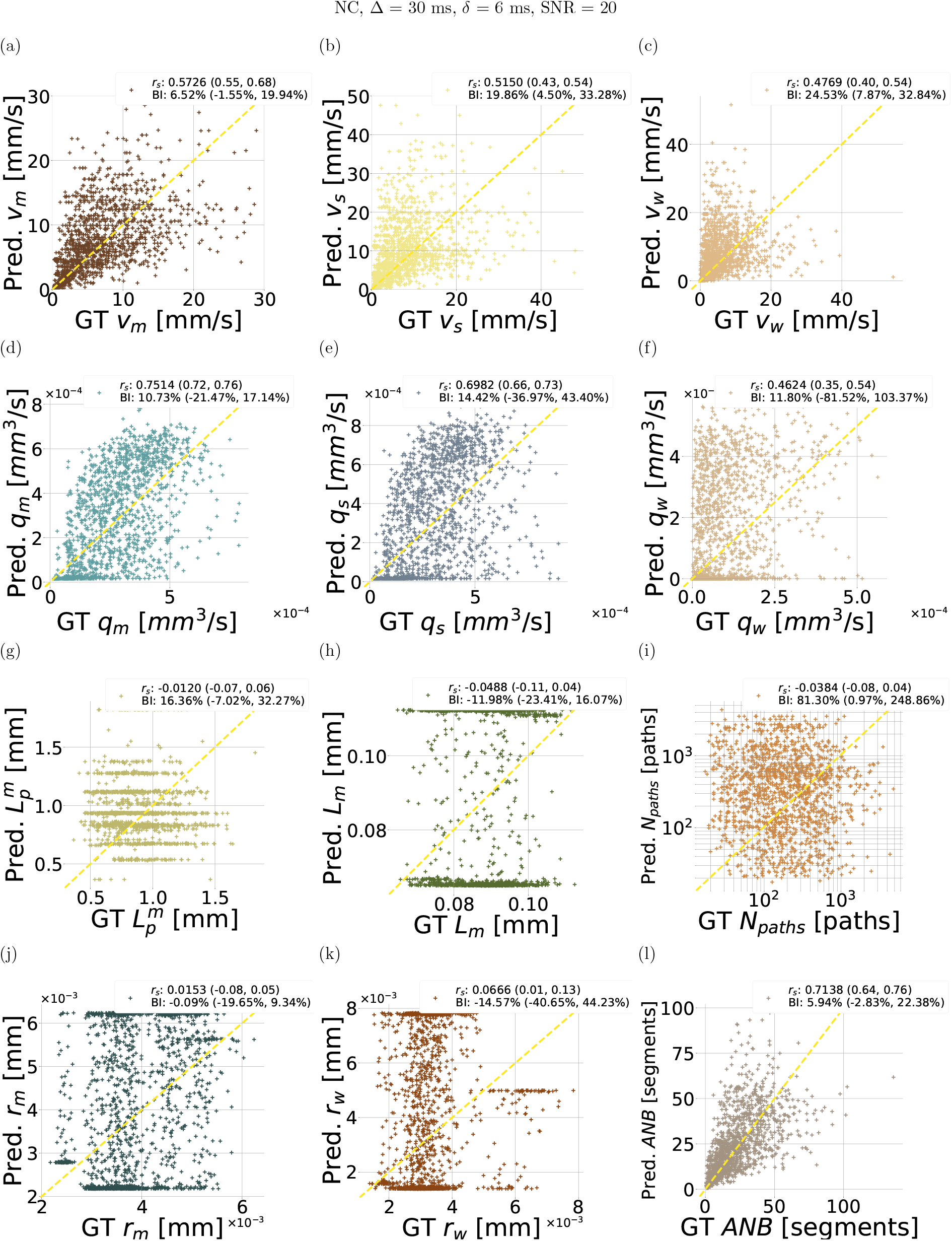
Scatter plots of estimated vascular parameters against ground truth values from the leave-one-out fitting procedure implemented *in silico*. The figure refers to the NC protocol, with Δ = 30 ms and *δ* = 6 ms, SNR = 20. From top to bottom: first row, mean velocity *v*_*m*_ in (a), standard deviation of velocity *v*_*s*_ in (b), path-weighted mean velocity *v*_*w*_ in (c); second row, mean volumetric flow rate (VFR) *q*_*m*_ in (d), standard deviation of VFR *q*_*s*_ in (e), path-weighted mean VFR *q*_*w*_ in (f); third row, mean input/output path length 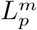 in (g), mean capillary segment length *L*_*m*_ in (h), number if input/output paths *N*_*paths*_ in (i); fourth row, mean capillary radius *r*_*m*_ in (j), path-weighted mean capillary radius *r*_*w*_ in (k), and apparent network branching *ANB* in (l). For each metric, the overall Spearman’s correlation coefficient *r*_*s*_ and Bias Index (BI) are reported, with the range of *r*_*s*_ and BI values obtained across leave-one-out folds. “GT” and “Pred.” respectively indicate ground truth and predicted metric values.

**Supporting Information Figure S6:**
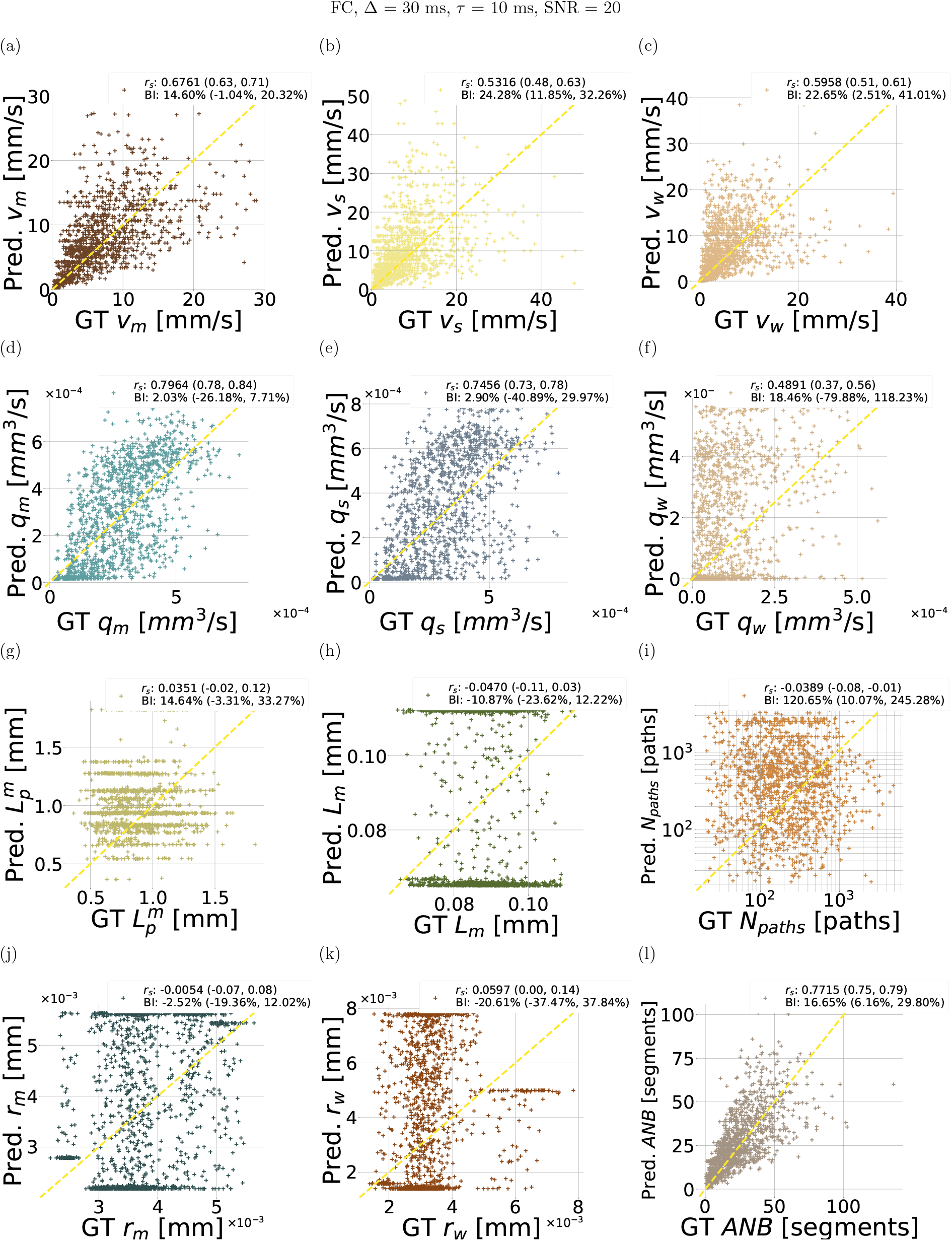
Scatter plots of estimated vascular parameters against ground truth values from the leave-one-out fitting procedure implemented *in silico*. The figure refers to the FC protocol, with Δ = 30 ms and *τ* = 10 ms, SNR = 20. From top to bottom: first row, mean velocity *v*_*m*_ in (a), standard deviation of velocity *v*_*s*_ in (b), path-weighted mean vel^1^ocity *v*_*w*_ in (c); second row, mean volumetric flow rate (VFR) *q*_*m*_ in (d), standard deviation of VFR *q*_*s*_ in (e), path-weighted mean VFR *q*_*w*_ in (f); third row, mean input/output path length 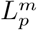 in (g), mean capillary segment length *L*_*m*_ in (h), number if input/output paths *N*_*paths*_ in (i); fourth row, mean capillary radius *r*_*m*_ in (j), path-weighted mean capillary radius *r*_*w*_ in (k), and apparent network branching *ANB* in (l). For each metric, the overall Spearman’s correlation coefficient *r*_*s*_ and Bias Index (BI) are reported, with the range of *r*_*s*_ and BI values obtained across leave-one-out folds. “GT” and “Pred.” respectively indicate ground truth and predicted metric values.

**Supporting Information Figure S7:**
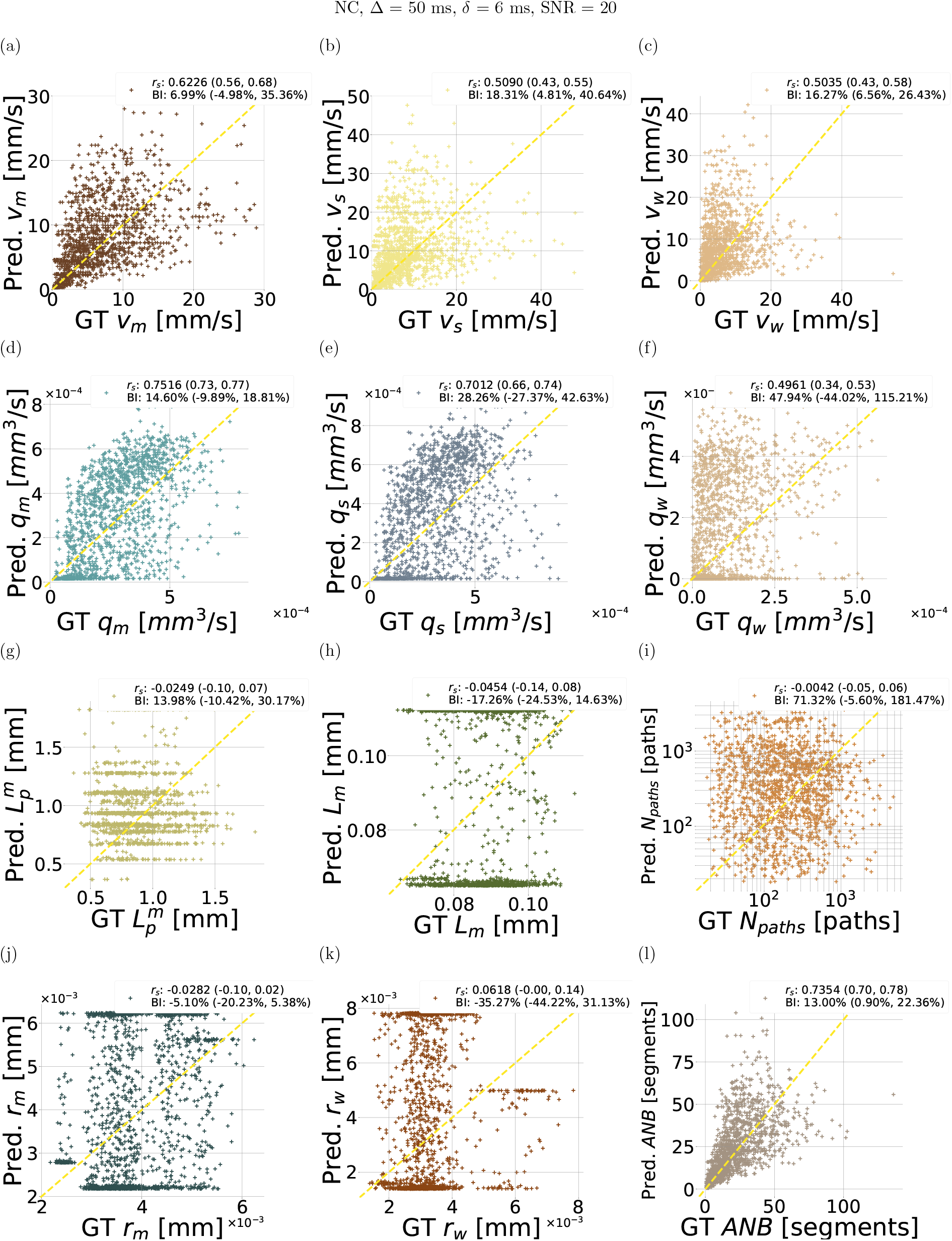
Scatter plots of estimated vascular parameters against ground truth values from the leave-one-out fitting procedure implemented *in silico*. The figure refers to the NC protocol, with Δ = 50 ms and *δ* = 6 ms, SNR = 20. From top to bottom: first row, mean velocity *v*_*m*_ in (a), standard deviation of velocity *v*_*s*_ in (b), path-weighted mean vel^1^ocity *v*_*w*_ in (c); second row, mean volumetric flow rate (VFR) *q*_*m*_ in (d), standard deviation of VFR *q*_*s*_ in (e), path-weighted mean VFR *q*_*w*_ in (f); third row, mean input/output path length 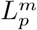 in (g), mean capillary segment length *L*_*m*_ in (h), number if input/output paths *N*_*paths*_ in (i); fourth row, mean capillary radius *r*_*m*_ in (j), path-weighted mean capillary radius *r*_*w*_ in (k), and apparent network branching *ANB* in (l). For each metric, the overall Spearman’s correlation coefficient *r*_*s*_ and Bias Index (BI) are reported, with the range of *r*_*s*_ and BI values obtained across leave-one-out folds. “GT” and “Pred.” respectively indicate ground truth and predicted metric values.

**Supporting Information Figure S8:**
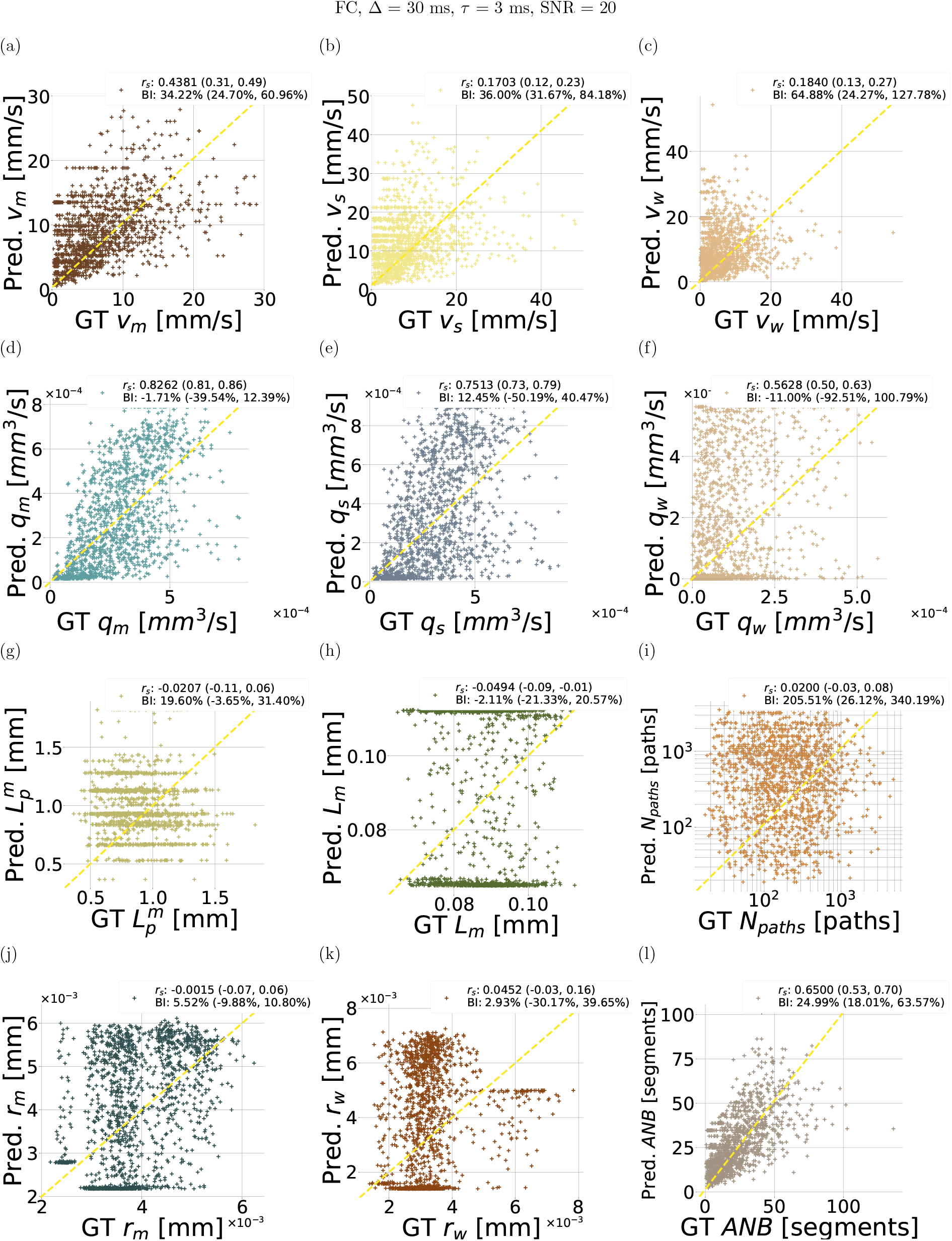
Scatter plots of estimated vascular parameters against ground truth values from the leave-one-out fitting procedure implemented *in silico*. The figure refers to the FC protocol, with Δ = 30 ms and *τ* = 3 ms, SNR = 20. From top to bottom: first row, mean velocity *v*_*m*_ in (a), standard deviation of velocity *v*_*s*_ in (b), path-weighted mean vel^1^ocity *v*_*w*_ in (c); second row, mean volumetric flow rate (VFR) *q*_*m*_ in (d), standard deviation of VFR *q*_*s*_ in (e), path-weighted mean VFR *q*_*w*_ in (f); third row, mean input/output path length 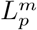 in (g), mean capillary segment length *L*_*m*_ in (h), number if input/output paths *N*_*paths*_ in (i); fourth row, mean capillary radius *r*_*m*_ in (j), path-weighted mean capillary radius *r*_*w*_ in (k), and apparent network branching *ANB* in (l). For each metric, the overall Spearman’s correlation coefficient *r*_*s*_ and Bias Index (BI) are reported, with the range of *r*_*s*_ and BI values obtained across leave-one-out folds. “GT” and “Pred.” respectively indicate ground truth and predicted metric values.

**Supporting Information Figure S9:**
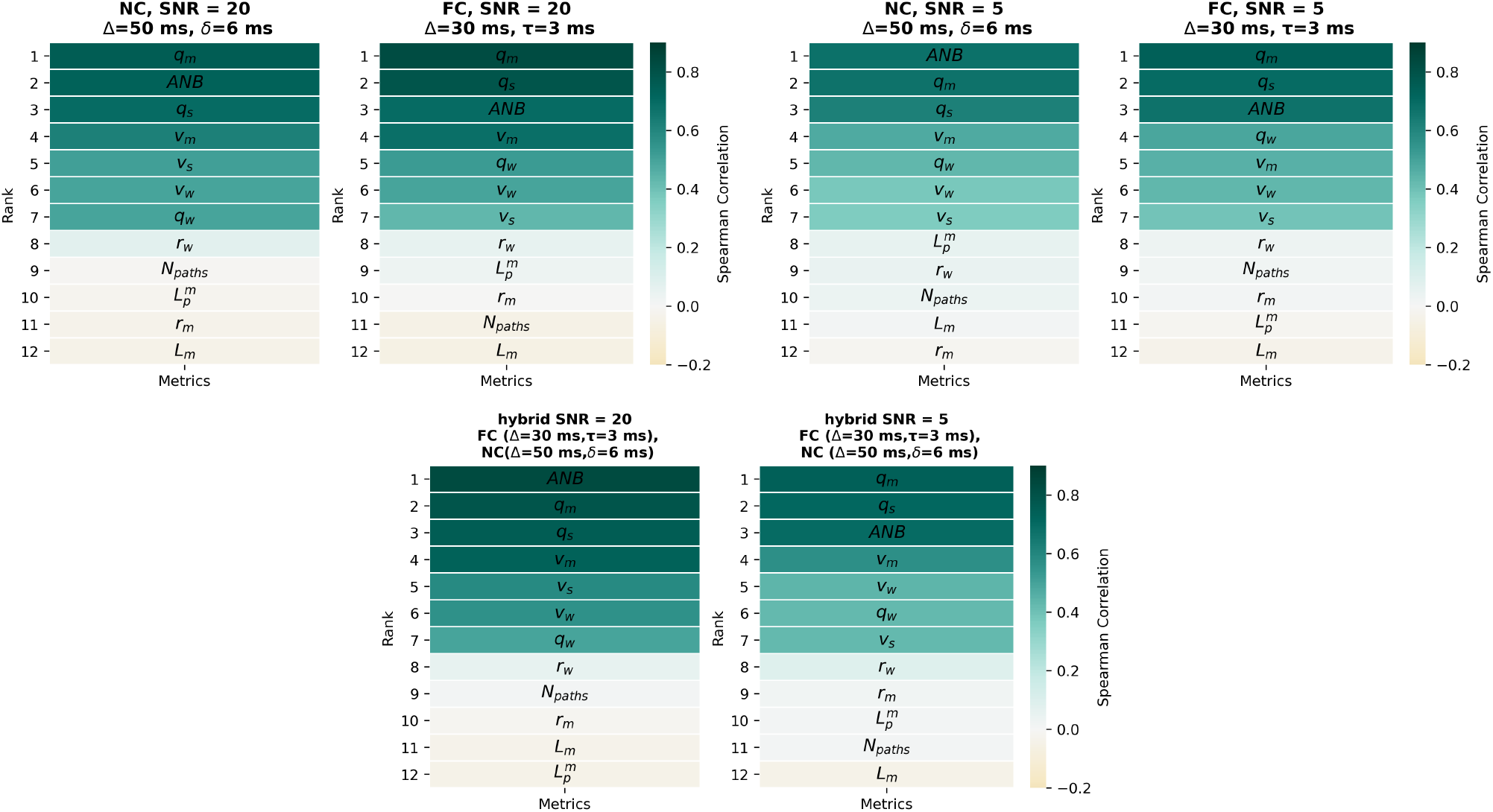
Ranking of microvascular metrics based on the goodness of the estimation, i.e., based on sorting the Spearman’s correlation *r*_*s*_ coefficients of all microvascular properties. For each property, *r*_*s*_ was computed by relating ground truth and estimated metric values. Ranking was obtained for the NC and FC protocols for SNR = 20 and SNR = 5, and for a hybrid protocol alternating measurements from the NC and the FC gradient waveforms. Top row, from left to right: ranking for the NC protocol (SNR = 20), FC protocol (SNR = 20), NC protocol (SNR = 5), FC protocol (SNR = 5). Bottom row, from left to right: ranking for the hybrid protocol at SNR = 20 and SNR = 5. The figure refers to the following diffusion time: Δ = 30 ms, *δ* = 6 ms for the NC protocol; Δ = 30 ms, *τ* = 10 ms for the FC protocol.

**Supporting Information Figure S10:**
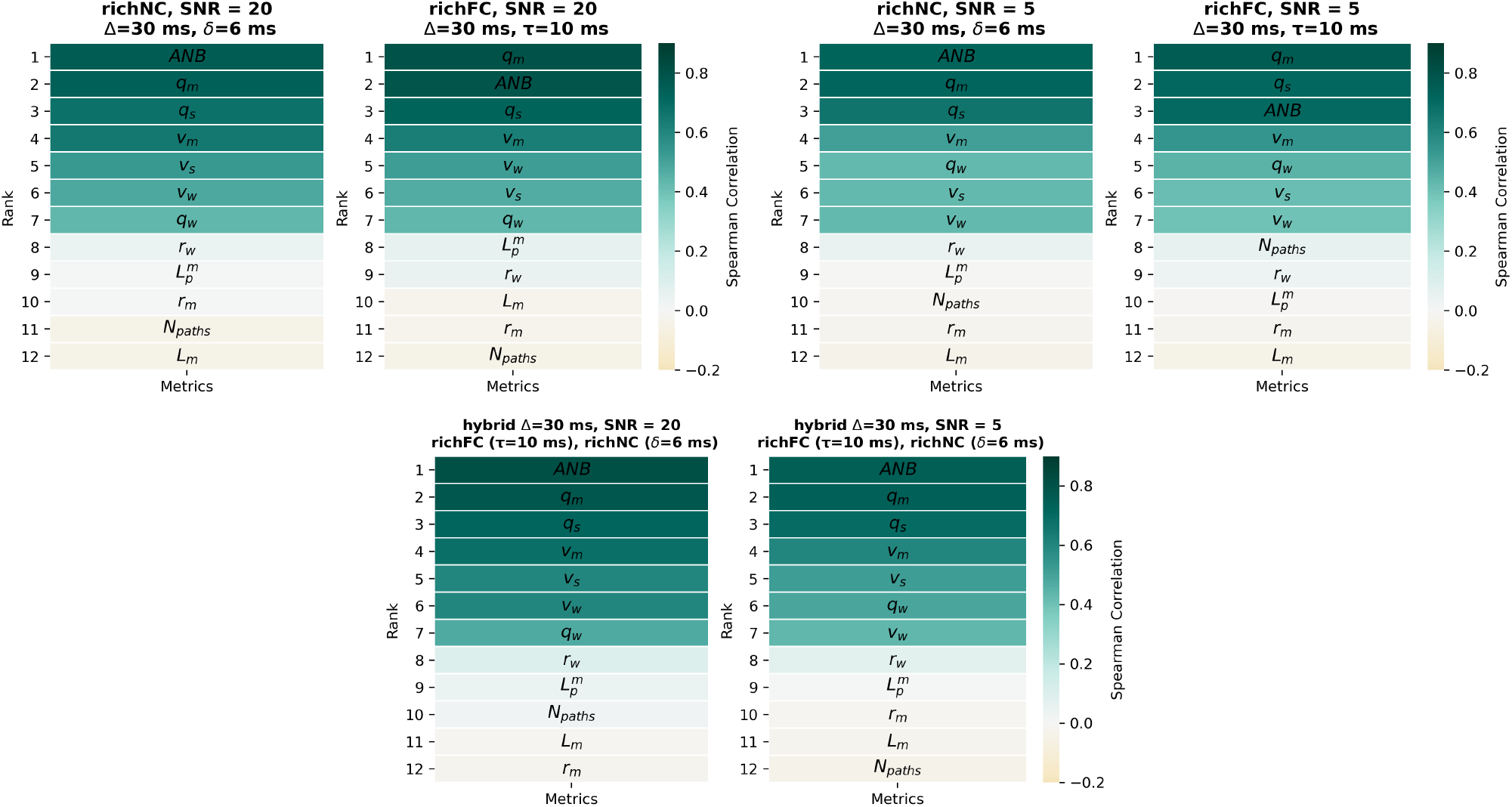
Ranking of microvascular metrics based on the goodness of the estimation, i.e., based on sorting the Spearman’s correlation *r*_*s*_ coefficients of all microvascular properties. For each property, *r*_*s*_ was computed by relating ground truth and estimated metric values. Ranking was obtained for the richNC and richFC protocols (with directional averaging) for SNR = 20 and SNR = 5, and for a hybrid protocol alternating measurements from the richNC and the richFC gradient waveforms. Top row, from left to right: ranking for the richNC protocol (SNR = 20), richFC protocol (SNR = 20), richNC protocol (SNR = 5), richFC protocol (SNR = 5). Bottom row, from left to right: ranking for the hybrid protocol at SNR = 20 and SNR = 5. The figure refers to the following diffusion time: Δ = 30 ms, *δ* = 6 ms for the NC protocol; Δ = 30 ms, *τ* = 10 ms for the FC protocol.

**Supporting Information Figure S11:**
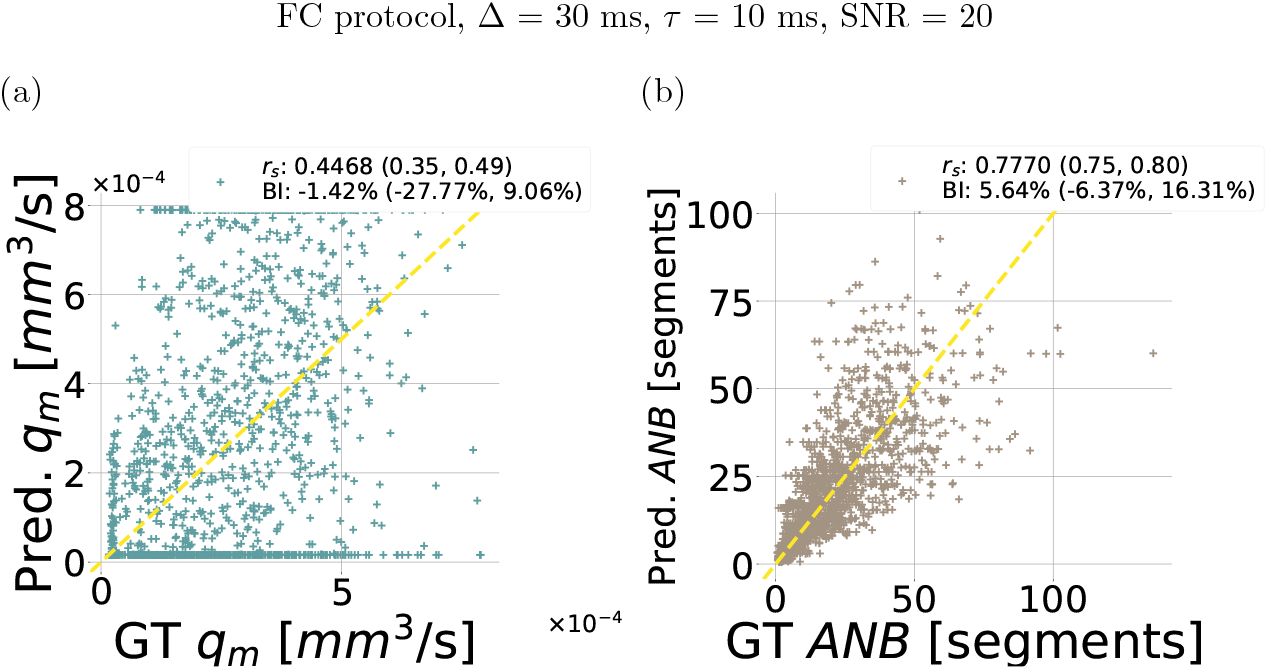
Example of quality of microvascular parameter fitting when two parameters are estimated jointly. The figure refers to the joint estimation of *q*_*m*_ and *ANB* for the flowcompensated (FC) protocol characterised by diffusion gradient timings of Δ = 30 ms and *τ* = 10 ms, for an SNR at *b* = 0 of 20.

**Supporting Information Figure S12:**
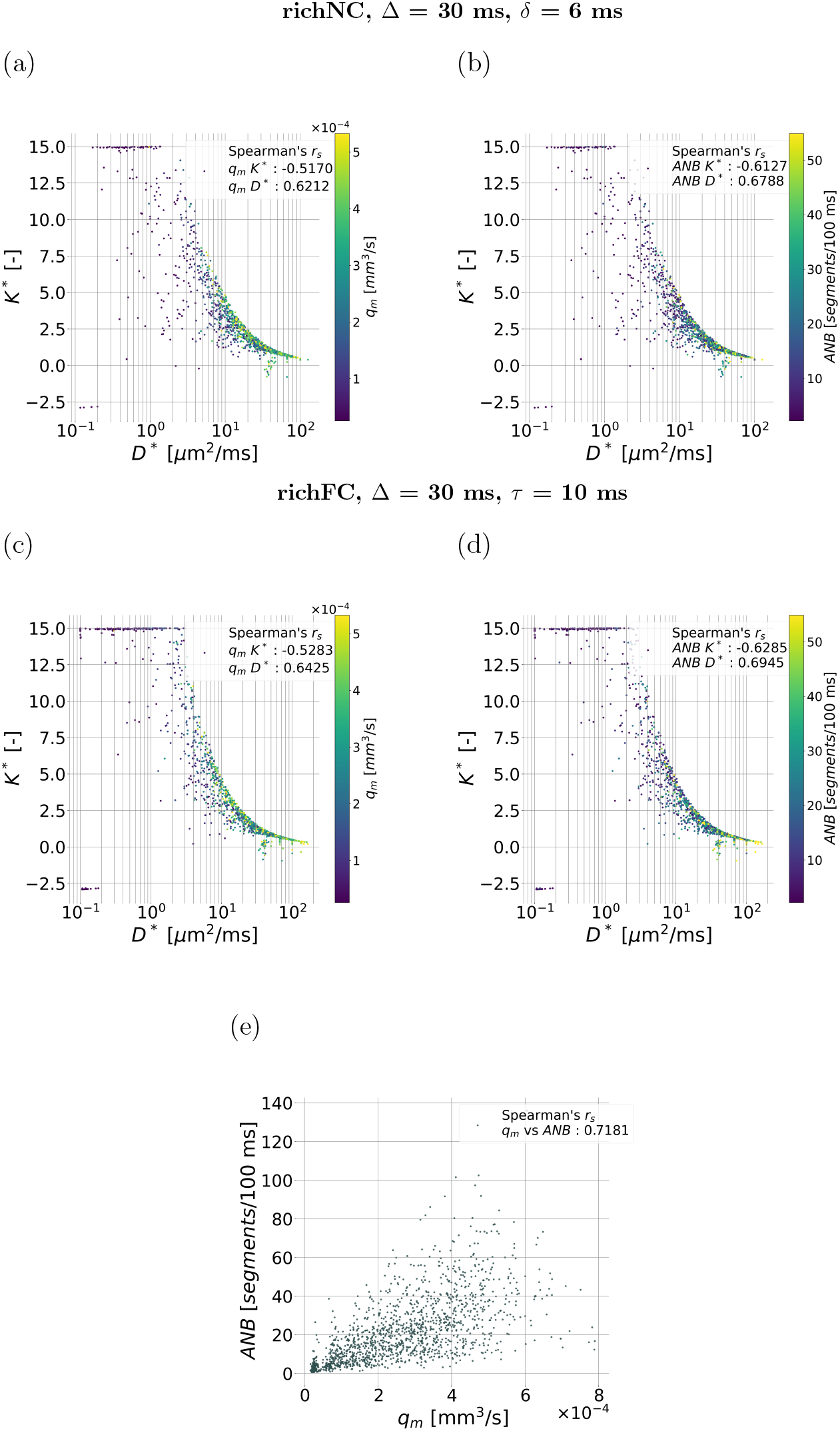
Scatter plots visualising the relationship between the vascular dMR signal cumulants and the top-ranking metrics selected from the *in silico* study. The figure scatters the apparent vascular diffusion and kurtosis coefficients (*D*^*^ and *K*^*^) against each other, colouring the points according to the mean VFR *q*_*m*_ and the apparent network branching *ANB*, thus visualising the dependence of these two metrics on *D*^*^ and *K*^*^, i.e., *q*_*m*_ = *f* (*D*^*^, *K*^*^) and *ANB* = *f* (*D*^*^, *K*^*^). Panels (a) and (b), top row: results for the richNC protocol (*q*_*m*_ = *f* (*D*^*^, *K*^*^) in (a); *ANB* = *f* (*D*^*^, *K*^*^) in (b); Spearman’s correlation coefficients between *q*_*m*_ and *D*^*^ and *K*^*^, and between *ANB* and *D*^1*^ and *K*^*^ are also reported). Panels (c) and (d), central row: results for the richFC protocol (*q*_*m*_ = *f* (*D*^*^, *K*^*^) in (c); *ANB* = *f* (*D*^*^, *K*^*^) in (d); Spearman’s correlation coefficients between *q*_*m*_ and *D*^*^ and *K*^*^, and between *ANB* and *D*^*^ and *K*^*^ are also reported). Panel (e), bottom row: scatter plot relating *q*_*m*_ and *ANB*, with corresponding Spearman’s correlation coefficient.

**Supporting Information Figure S13:**
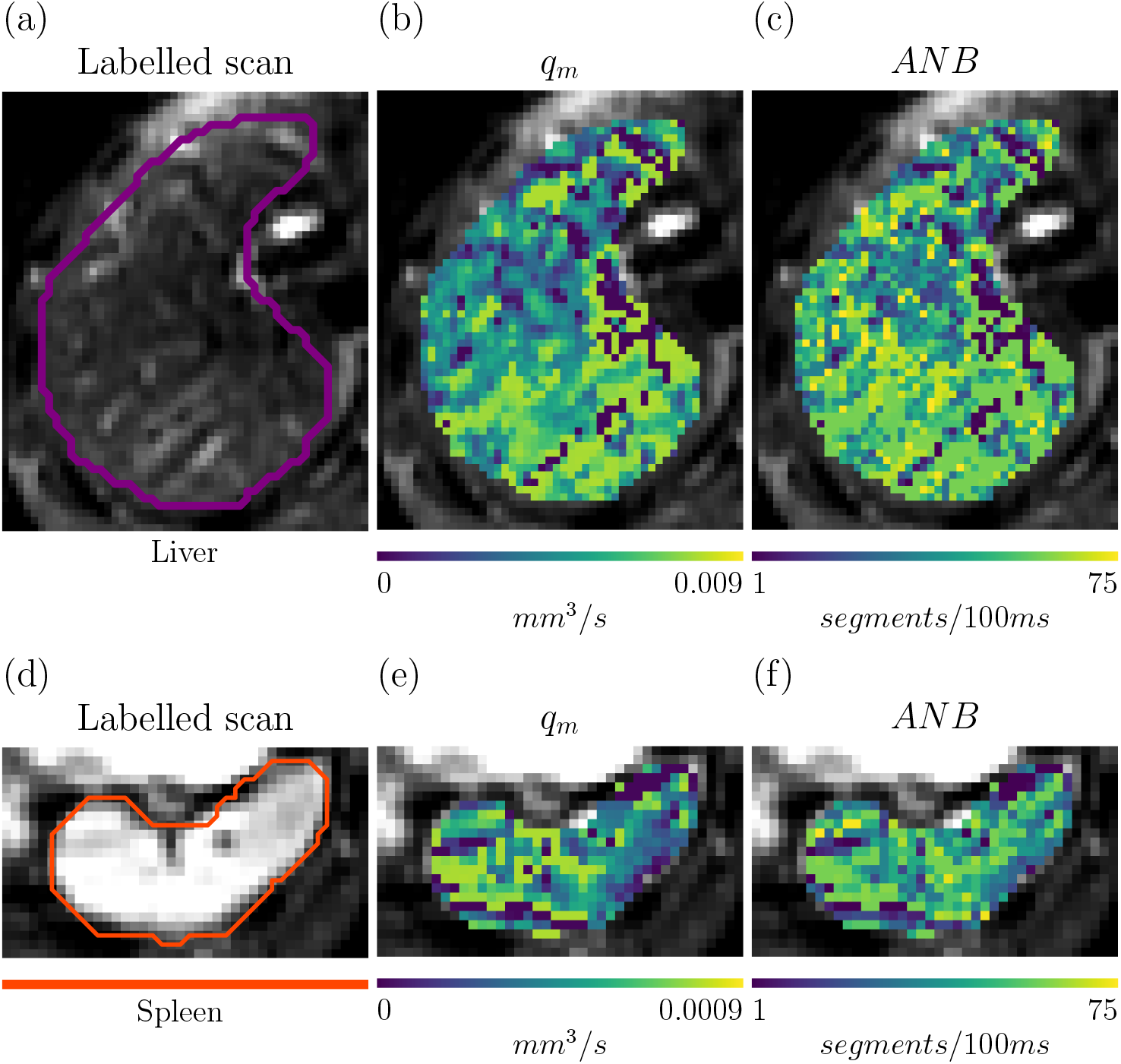
Microvascular parameter estimation in a healthy male volunteer *in vivo*. Top: microvascular parameter mapping in the liver parenchyma. From left to right, (a): structural, anatomical T2-weighted scan of the liver; (b) *q*_*m*_ map in the liver; (c): *ANB* map in the liver. Bottom: microvascular parameter mapping in the spleen. From left to right, (d): structural, anatomical T2-weighted scan of the spleen; (e) *q*_*m*_ map in the spleen; (f): *ANB* map in the spleen.

## References

[1] V. G. Kiselev, “Fundamentals of diffusion mri physics,” NMR in Biomedicine, vol. 30, no. 3, p. e3602, 2017.

[2] C. Ahn, S. Lee, O. Nalcioglu, and Z. Cho, “The effects of random directional distributed flow in nuclear magnetic resonance imaging,” Medical physics, vol. 14, no. 1, pp. 43–48, 1987.

[3] D. Le Bihan, M. Iima, C. Federau, and E. E. Sigmund, Intravoxel incoherent motion (IVIM) MRI: principles and applications. CRC Press, 2018.

[4] D. Le Bihan, “What can we see with ivim mri?,” Neuroimage, vol. 187, pp. 56–67, 2019.

[5] D. Le Bihan, E. Breton, D. Lallemand, P. Grenier, E. Cabanis, and M. Laval-Jeantet, “Mr imaging of intravoxel incoherent motions: application to diffusion and perfusion in neurologic disorders.,” Radiology, vol. 161, no. 2, pp. 401–407, 1986.

[6] E. K. Englund, D. A. Reiter, B. Shahidi, and E. E. Sigmund, “Intravoxel incoherent motion magnetic resonance imaging in skeletal muscle: review and future directions,” Journal of Magnetic Resonance Imaging, vol. 55, no. 4, pp. 988–1012, 2022.

[7] A. Arian, F. Z. Seyed-Kolbadi, S. Yaghoobpoor, H. Ghorani, A. Saghazadeh, and D. J. Ghadimi, “Diagnostic accuracy of intravoxel incoherent motion (ivim) and dynamic contrast-enhanced (dce) mri to differentiate benign from malignant breast lesions: A systematic review and meta-analysis,” European Journal of Radiology, vol. 167, p. 111051, 2023.

[8] A. Ahlgren, L. Knutsson, R. Wirestam, M. Nilsson, F. Ståhlberg, D. Topgaard, and S. Lasič, “Quantification of microcirculatory parameters by joint analysis of flow-compensated and non-flow-compensated intravoxel incoherent motion (ivim) data,” NMR in Biomedicine, vol. 29, no. 5, pp. 640–649, 2016.

[9] L. A. Scott, B. R. Dickie, S. D. Rawson, G. Coutts, T. L. Burnett, S. M. Allan, G. J. Parker, and L. M. Parkes, “Characterisation of microvessel blood velocity and segment length in the brain using multidiffusion-time diffusion-weighted mri,” Journal of Cerebral Blood Flow & Metabolism, vol. 41, no. 8, pp. 1939–1953, 2021.

[10] A. Luciani, A. Vignaud, M. Cavet, J. Tran Van Nhieu, A. Mallat, L. Ruel, A. Laurent, J.-F. Deux, P. Brugieres, and A. Rahmouni, “Liver cirrhosis: intravoxel incoherent motion mr imaging—pilot study,” Radiology, vol. 249, no. 3, pp. 891–899, 2008.

[11] J. H. Yoon, J. M. Lee, M. H. Yu, B. Kiefer, J. K. Han, and B. I. Choi, “Evaluation of hepatic focal lesions using diffusion-weighted mr imaging: comparison of apparent diffusion coefficient and intravoxel incoherent motion-derived parameters,” Journal of Magnetic Resonance Imaging, vol. 39, no. 2, pp. 276–285, 2014.

[12] J. Tao, Z. Yin, X. Li, Y. Zhang, K. Zhang, Y. Yang, S. Fang, and S. Wang, “Correlation between ivim parameters and microvessel architecture: Direct comparison of mri images and pathological slices in an orthotopic murine model of rhabdomyosarcoma,” European Radiology, vol. 33, no. 12, pp. 8576–8584, 2023.

[13] E. Fokkinga, J. A. Hernandez-Tamames, A. Ianus, M. Nilsson, C. M. Tax, R. Perez-Lopez, and F. Grussu, “Advanced diffusion-weighted mri for cancer microstructure assessment in body imaging, and its relationship with histology,” Journal of Magnetic Resonance Imaging, vol. 60, no. 4, pp. 1278–1304, 2024.

[14] D. Wu and J. Zhang, “Evidence of the diffusion time dependence of intravoxel incoherent motion in the brain,” Magnetic resonance in medicine, vol. 82, no. 6, pp. 2225–2235, 2019.

[15] G. Fournet, J.-R. Li, A. M. Cerjanic, B. P. Sutton, L. Ciobanu, and D. Le Bihan, “A two-pool model to describe the ivim cerebral perfusion,” Journal of Cerebral Blood Flow & Metabolism, vol. 37, no. 8, pp. 2987–3000, 2017.

[16] G. Simchick, R. Geng, Y. Zhang, and D. Hernando, “b value and first-order motion moment optimized data acquisition for repeatable quantitative intravoxel incoherent motion dwi,” Magnetic resonance in medicine, vol. 87, no. 6, pp. 2724–2740, 2022.

[17] G. Simchick and D. Hernando, “Precision of region of interest-based tri-exponential intravoxel incoherent motion quantification and the role of the intervoxel spatial distribution of flow velocities,” Magnetic resonance in medicine, vol. 88, no. 6, pp. 2662–2678, 2022.

[18] T. Führes, A. J. Riexinger, M. Loh, J. Martin, A. Wetscherek, T. A. Kuder, M. Uder, B. Hensel, and F. B. Laun, “Echo time dependence of biex-ponential and triexponential intravoxel incoherent motion parameters in the liver,” Magnetic resonance in medicine, vol. 87, no. 2, pp. 859–871, 2022.

[19] V. P. Van, F. Schmid, G. Spinner, S. Kozerke, and C. Federau, “Simulation of intravoxel incoherent perfusion signal using a realistic capillary network of a mouse brain,” NMR in Biomedicine, vol. 34, no. 7, p. e4528, 2021.

[20] J. Weine, C. McGrath, P. Dirix, S. Buoso, and S. Kozerke, “Cmrsim–a python package for cardiovascular mr simulations incorporating complex motion and flow,” Magnetic Resonance in Medicine, vol. 91, no. 6, pp. 2621–2637, 2024.

[21] M. Lashgari, Z. Yang, M. O. Bernabeu, J.-R. Li, and A. F. Frangi, “Spindoctor-ivim: A virtual imaging framework for intravoxel incoherent motion mri,” Medical Image Analysis, vol. 99, p. 103369, 2025.

[22] A. K. Voronova, A. Grigoriou, K. Bernatowicz, S. Simonetti, G. Serna, N. Roson, M. Escobar, M. Vieito, P. Nuciforo, R. Toledo, et al., “Spinflowsim: A blood flow simulation framework for histology-informed diffusion mri microvasculature mapping in cancer,” Medical image analysis, vol. 102, p. 103531, 2025.

[23] A. Wetscherek, B. Stieltjes, and F. B. Laun, “Flow-compensated intravoxel incoherent motion diffusion imaging,” Magnetic resonance in medicine, vol. 74, no. 2, pp. 410–419, 2015.

[24] E. Fieremans and H.-H. Lee, “Physical and numerical phantoms for the validation of brain microstructural mri: A cookbook,” Neuroimage, vol. 182, pp. 39–61, 2018.

[25] E. O. Stejskal and J. E. Tanner, “Spin diffusion measurements: spin echoes in the presence of a time-dependent field gradient,” The journal of chemical physics, vol. 42, no. 1, pp. 288–292, 1965.

[26] O. J. Gurney-Champion, S. S. Rauh, K. Harrington, U. Oelfke, F. B. Laun, and A. Wetscherek, “Optimal acquisition scheme for flow-compensated intravoxel incoherent motion diffusion-weighted imaging in the abdomen: An accurate and precise clinically feasible protocol,” Magnetic resonance in medicine, vol. 83, no. 3, pp. 1003–1015, 2020.

[27] C.-F. Westin, H. Knutsson, O. Pasternak, F. Szczepankiewicz, E. Özarslan, D. van Westen, C. Mattisson, M. Bogren, L. J. O’donnell, M. Kubicki, et al., “Q-space trajectory imaging for multidimensional diffusion mri of the human brain,” Neuroimage, vol. 135, pp. 345–362, 2016.

[28] E. Caruyer, C. Lenglet, G. Sapiro, and R. Deriche, “Design of multishell sampling schemes with uniform coverage in diffusion mri,” Magnetic resonance in medicine, vol. 69, no. 6, pp. 1534–1540, 2013.

[29] J. Veraart, E. Fieremans, and D. S. Novikov, “Diffusion mri noise mapping using random matrix theory,” Magnetic resonance in medicine, vol. 76, no. 5, pp. 1582–1593, 2016.

[30] J. Veraart, D. S. Novikov, D. Christiaens, B. Ades-Aron, J. Sijbers, and E. Fieremans, “Denoising of diffusion mri using random matrix theory,” Neuroimage, vol. 142, pp. 394–406, 2016.

[31] E. Panagiotaki, T. Schneider, B. Siow, M. G. Hall, M. F. Lythgoe, and D. C. Alexander, “Compartment models of the diffusion mr signal in brain white matter: a taxonomy and comparison,” Neuroimage, vol. 59, no. 3, pp. 2241–2254, 2012.

[32] C. Macarro, K. Bernatowicz, A. Garcia-Ruiz, G. Serna, C. Monreal-Agüero, S. Simonetti, M. Figini, J. F. Corral, V. Garay, M. Vidorreta, et al., “Enhancing tumor microstructural quantification with machine learning and diffusion-relaxation mri,” Journal of Magnetic Resonance Imaging, vol. 61, no. 2, p. 1018, 2025.

[33] O. J. Gurney-Champion, R. Klaassen, M. Froeling, S. Barbieri, J. Stoker, M. R. Engelbrecht, J. W. Wilmink, M. G. Besselink, A. Bel, H. W. Van Laarhoven, et al., “Comparison of six fit algorithms for the intra-voxel incoherent motion model of diffusion-weighted magnetic resonance imaging data of pancreatic cancer patients,” PloS one, vol. 13, no. 4, p. e0194590, 2018.

[34] Y. Cui, H. Dyvorne, C. Besa, N. Cooper, and B. Taouli, “Ivim diffusion-weighted imaging of the liver at 3.0 t: Comparison with 1.5 t,” European journal of radiology open, vol. 2, pp. 123–128, 2015.

[35] P. J. Basser, J. Mattiello, and D. LeBihan, “Mr diffusion tensor spectroscopy and imaging,” Bio-physical journal, vol. 66, no. 1, pp. 259–267, 1994.

[36] M. Notohamiprodjo, H. Chandarana, A. Mikheev, H. Rusinek, J. Grinstead, T. Feiweier, J. G. Raya, V. S. Lee, and E. E. Sigmund, “Combined intravoxel incoherent motion and diffusion tensor imaging of renal diffusion and flow anisotropy,” Magnetic resonance in medicine, vol. 73, no. 4, pp. 1526–1532, 2015.

[37] F. Hilbert, M. Bock, H. Neubauer, S. Veldhoen, T. Wech, T. A. Bley, and H. Köstler, “An intravoxel oriented flow model for diffusion-weighted imaging of the kidney,” NMR in Biomedicine, vol. 29, no. 10, pp. 1403–1413, 2016.

[38] A. Finkelstein, X. Cao, C. Liao, G. Schifitto, and J. Zhong, “Diffusion encoding methods in mri: perspectives and challenges,” Investigative Magnetic Resonance Imaging, vol. 26, no. 4, pp. 208–219, 2022.

[39] G. Simchick and D. Hernando, “Optimized acquisition for simultaneous intravoxel incoherent motion and r2 quantification in the liver,” Magnetic Resonance in Medicine, vol. (early view), 2025.

[40] P. Feldman, M. Fainstein, V. Siless, C. Delrieux, and E. Iarussi, “Recursive variational autoen-coders for 3d blood vessel generative modeling,” Medical Image Analysis, p. 103703, 2025.

[41] P. Blinder, P. S. Tsai, J. P. Kaufhold, P. M. Knutsen, H. Suhl, and D. Kleinfeld, “The cortical angiome: an interconnected vascular network with noncolumnar patterns of blood flow,” Nature neuroscience, vol. 16, no. 7, pp. 889–897, 2013.

[42] V. P. Van, F. Schmid, G. Spinner, S. Kozerke, and C. Federau, “Simulation of intravoxel incoherent perfusion signal using a realistic capillary network of a mouse brain,” NMR in Biomedicine, vol. 34, no. 7, p. e4528, 2021.

[43] D. C. Alexander, “A general framework for experiment design in diffusion mri and its application in measuring direct tissue-microstructure features,” Magnetic Resonance in Medicine, vol. 60, no. 2, pp. 439–448, 2008.

[44] S. Kyriazi, M. Blackledge, D. J. Collins, and N. M. Desouza, “Optimising diffusion-weighted imaging in the abdomen and pelvis: comparison of image quality between monopolar and bipolar single-shot spin-echo echo-planar sequences,” European radiology, vol. 20, no. 10, pp. 2422–2431, 2010.

